# Diagnostic Efficacy of Various Imaging Modalities Across Different Stages of Prostate Cancer: A Network Meta-Analysis of Diagnostic Studies

**DOI:** 10.1101/2024.09.28.24314285

**Authors:** Chengdong Shi, Kai Yu, Yu Hu, Yuantao Wang, Fan Bu, Ji Lu, Weigang Wang

## Abstract

**Purpose:** To assess the diagnostic performance of various imaging modalities in detecting and monitoring prostate cancer across different disease stages using diagnostic test accuracy (DTA) and network meta-analysis (NMA).

**Methods:** A systematic literature review was conducted to identify studies evaluating mpMRI, PSMA PET/CT, MRE, MRSI, BS, CT, PET, and other tracers for prostate cancer detection. Data on sensitivity, specificity, PPV, NPV, and detection rate were extracted and analyzed using NMA.

**Result:** Across 123 studies involving 9,371 patients, 68Ga-P16-093 PET/CT and 68Ga-PSMA-617 PET/CT showed high diagnostic accuracy in early-phase prostate cancer. For lymph node metastasis, 68Ga-PSMA-11 PET/MRI was the most sensitive. 18F-DCFPyL PET/CT had the highest specificity and PPV, while 18F-PSMA-1007 PET/CT had the highest NPV. In bone metastasis, 18F-PSMA-1007 PET/MRI excelled in sensitivity and NPV, while 18F-Fluciclovine PET/CT had the highest specificity and PPV. For biochemical recurrence, 18F-PSMA-1007 PET/CT had the highest lesion detection rate, and for different radiotracers, 18F-PSMA-1007 had the highest detection rate.

**Conclusion:** This network meta-analysis comprehensively evaluated the diagnostic efficacy of various imaging modalities for prostate cancer across different stages. Our findings underscore the strengths and limitations of each imaging technique in detecting and staging prostate cancer.

## 1. Introduction

Prostate cancer is one of the most common malignant tumors of the male genitourinary system. According to the Global Cancer Statistics 2022 report, there were approximately 1.46 million new cases of prostate cancer in 2022, accounting for 14.2% of new cancer cases in men, making it the second most common malignancy in men after lung cancer. Additionally, there were about 390,000 new deaths due to prostate cancer, representing 7.3% of cancer-related deaths in men, ranking it fifth overall (1). Age has been identified as an independent risk factor for prostate cancer incidence (2). With the continuous growth and aging of the population, the incidence of prostate cancer is increasing annually, posing a serious threat to male health and presenting greater challenges and burdens to global public health systems (3, 4). Prostate cancer tends to exhibit subtle clinical manifestations in its early stages, leading many patients to seek medical attention only after the tumor has infiltrated or metastasized, significantly increasing mortality rates. In recent years, advancements in serology, imaging techniques, and surgical methods have contributed to higher detection rates and treatment success rates (5, 6). Among these, imaging examinations play a crucial role in screening, early diagnosis, tumor staging, surgical selection, and predicting the recurrence prognosis of prostate cancer (7, 8).

Currently, the main screening methods for prostate cancer are digital rectal examination (DRE) and serum prostate-specific antigen (PSA) testing. The introduction of PSA testing has significantly increased the detection rate of prostate cancer. However, there is an ongoing debate about its contribution to the overdiagnosis and overtreatment of clinically insignificant tumors (9, 10). DRE has its limitations, particularly in detecting tumors located deep within the prostate or those that are smaller in size. A meta-analysis indicates that the sensitivity and specificity of DRE performed by primary care physicians are 0.51 and 0.59 (11). Therefore, the overuse of PSA testing and the low specificity and sensitivity of DRE necessitate the use of additional diagnostic methods to enhance the accuracy of prostate cancer screening. To improve tumor detection rates and achieve more accurate staging, the application of multiparametric magnetic resonance imaging (mpMRI) plays a crucial role in optimizing clinical diagnosis and biopsy procedures(12). MpMRI assesses the likelihood of tumor presence based on the Prostate Imaging Reporting and Data System (PI-RADS) and can enhance biopsy efficacy through visual guidance. A meta-analysis showed that when PI-RADS is correctly used, the pooled sensitivity for diagnosing prostate cancer is ([OR] 0.82, 95% CI [0.72-0.89]) and specificity is ([OR] 0.82, 95% CI [0.67-0.92])(13). Additionally, a study by Veeru demonstrated that compared to systematic biopsy methods, MRI-targeted biopsy (MRI-TB) has a higher detection rate for clinically significant prostate cancer ([OR] 1.16, 95% CI [1.09-1.24]) while identifying fewer cases of clinically insignificant prostate cancer (14). However, study data also show that the positive predictive value of suspicious mpMRI for clinically significant prostate cancer (csPCa) is about 40%-50%, and the negative predictive value is about 80%-90%, indicating that more than half of patients with positive mpMRI results underwent unnecessary prostate biopsies or overtreatment. Additionally, the diagnostic sensitivity and specificity of mpMRI decrease for patients with a PI-RADS score of 3 (15, 16).

Other traditional imaging techniques, such as computed tomography (CT) and bone scintigraphy (BS), are still widely used in clinical practice, but each has its limitations. CT has low sensitivity for the initial diagnosis of prostate cancer and offers a single imaging mode, while BS is primarily used to assess bone metastasis in prostate cancer patients. Additionally, imaging modalities like magnetic resonance spectroscopy imaging (MRSI) and magnetic resonance elastography (MRE) are gradually maturing. However, these imaging techniques are generally used as auxiliary and supplementary diagnostic tools, and their cost-effectiveness and diagnostic efficiency still require further research (17).

The advent of positron emission tomography (PET) has provided a new direction for the diagnosis and treatment of prostate cancer. Initially, PET imaging used the 18F-FDG (Fluoro-2-deoxy-D-glucose) tracer, which has high sensitivity and can perform whole-body scans to detect small lesions and metastases throughout the body. Subsequently, the introduction of new tracers, such as 18F-NaF and 11C-choline, further enhanced PET’s detection capabilities (18, 19). In recent years, prostate-specific membrane antigen (PSMA) PET has gained widespread recognition for its high sensitivity and specificity in diagnosing prostate cancer and assessing biochemical recurrence. PSMA is a membrane protein significantly overexpressed in prostate cancer tissue, with expression levels 100-1000 times higher than in normal prostate tissue, making it an ideal target for molecular imaging. Increasing evidence suggests that PSMA-PET/CT and PSMA-PET/MRI have higher diagnostic efficacy than traditional imaging techniques. The diversity of PSMA ligands also provides options for addressing individual differences in prostate cancer cases. Currently, the most commonly used ligands are 68Ga-PSMA-11, 18F-DCFPyL (Piflufolastat), and 18F-PSMA-1007 (20). Meanwhile, it has been suggested that PSMA PET/CT may reduce the economic burden of high-risk patients through higher accuracy efficacy (21). However, PSMA PET is not without its limitations. Some studies indicate that while PSMA PET/CT has high specificity, its sensitivity can be inconsistent. Additionally, different tracers used in PSMA PET have varying diagnostic efficacies, and their absorption rates in different body parts can vary, raising concerns about abnormal uptake in normal or benign lesions leading to overdiagnosis and overtreatment (22). For PSMA PET/MRI, although it offers clearer contrast for soft tissues and studies have shown it to be more effective in diagnosing primary prostate cancer and biochemical recurrence (BCR) compared to mpMRI, its diagnostic performance compared to PSMA PET/CT remains unclear (23, 24) Moreover, PSMA PET/MRI is more expensive, has longer examination times, and is more challenging to implement in primary care settings.

Currently, there is controversy regarding the diagnostic performance of PSMA-PET compared to other imaging modalities and among different ligands at various stages of prostate cancer. This study aims to evaluate the diagnostic effectiveness of different imaging methods in detecting and monitoring prostate cancer at different stages by analyzing existing literature data.

## 2. Method

### 2.1 Protocol

This systematic review and meta-analysis was conducted according to the Preferred Reporting Items for Systematic Reviews and Meta-analyses (PRISMA) of diagnostic test accuracy (DTA) studies(25). The study protocol was registered on the International Prospective Register of Systematic Reviews (PROSPERO; registration ID CRD42021248896).

The aim of this systematic review is to assess the diagnostic efficacy of different imaging methods in different stages of prostate cancer. The proposed systematic review aims to answer the following questions:

i. Which imaging modality has higher diagnostic performance in the early stages of prostate cancer?
ii. Which imaging method has greater accuracy in diagnosing lymph nodes and bone metastases in prostate cancer?
iii. Which method has a higher lesion detection rate for biochemical recurrence of prostate cancer?
iv. How does the diagnostic performance of combined imaging modalities compare to individual imaging methods at different stages of prostate cancer?

### 2.2. Literature search

PubMed, Cochrane, and Embase were searched to identify reports published up to June 30, 2024, addressing the diagnostic value of prostate cancer at different stages (early-stage prostate cancer, advanced-stage prostate cancer, metastatic prostate cancer including bone and lymph node metastasis, recurrent prostate cancer such as biochemical recurrent prostate cancer). The keywords used in our search strategy are reported in Table 1. Initial screening was performed independently by two investigators (Shi CD and Yu K) based on the titles and abstracts of the articles to identify ineligible reports. Reasons for exclusions were noted. Potentially relevant reports were subjected to a full-text review, and the relevance of the reports was confirmed after the data extraction process. Any discrepancies during the primary and secondary literature screening were resolved by referring to the senior authors (Wang WG and Lu J).

**Table 1:** Keywords used in the literature search strategy. This table lists the keywords used in the literature search to identify relevant studies for the systematic review and meta-analysis.

Primary endpoint: To assess the diagnostic value of various examination methods for prostate cancer at different stages (including early-stage, advanced-stage, metastatic prostate cancer such as bone and lymph node metastasis, recurrent prostate cancer such as biochemical recurrent prostate cancer) using Diagnostic Test Accuracy meta-analysis. Since most of the patients with biochemical recurrence of prostate cancer could not be verified as true-positive or true-negative by pathology or follow-up investigations, the present study was conducted to compare the rate of lesion detection by different examination modalities in the biochemical recurrence section.

Secondary endpoint: To further evaluate the specific diagnostic value of these examination methods in different stages of prostate cancer, including early-stage, advanced-stage, metastatic prostate cancer, and recurrent prostate cancer, and to compare their sensitivity and specificity in detecting prostate cancer at specific stages.

### 2.3. Inclusion and exclusion criteria

The population, intervention, control, and outcomes (PICO) for this study were determined by the co-authors as follows: Inclusion criteria: ① The content of the article is designed as a diagnostic experimental study. ② Studies need to compare the diagnostic effectiveness of two or more imaging examinations on the same patient group (mpMRI, PSMA PET, MRSI, CT, BS, PET). ③ A study that includes histopathological results or obtains pathological evidence based on clinical data, other examinations, or follow-up results. ④ Studies that include results such as true positive (TP), true negative (TN), false positive (FP), false negative (FN), sensitivity (Se), specificity (Sp), positive predictive value (PPV), negative predictive value (NPV) and detection rate (DR) or studies that can derive these results through other data calculations. If incomplete data existed for TP, TN, FP, FN, NPV, or PPV, they were calculated using the known variables Se and Sp to complete the data.

Exclusion criteria included: ① Duplicate literature; ② Non-original content such as reviews, letters to the editor, editorials, research protocols, case reports, brief communications, guidelines, and studies using other non-standard imaging modalities; ③ Studies with a sample size of fewer than ten individuals or lacking original data; ④ Non-English articles. We excluded all studies that did not evaluate the diagnostic accuracy of prostate cancer assessment in comparison to the reference method, which refers to the established standard diagnostic procedure for prostate cancer. This study focused solely on original research investigating the diagnostic accuracy of prostate biopsy, histopathological examination, and follow-up content for reported cancers, which are invasive procedures commonly used for prostate cancer diagnosis. Additionally, we excluded articles not published in English and scanned the references of all included papers for additional relevant studies.

### 2.4 Data extraction

Two investigators independently extracted the following information from the included articles: author, publication year, patient number, tumor stage and grade, as well as Se, Sp, and the counts of TP, FP, FN, TN, or DR for the primary outcomes. Discrepancies were resolved by consensus with coauthors.

### 2.5 Risk of Bias Assessment

The risk of bias in the included studies was evaluated using the revised Quality Assessment of Diagnostic Accuracy Studies tool (QUADAS-2) (26). The index test was defined as the value of Multi-Stage Prostate Cancer detection using various imaging modalities, with prostate needle biopsy and histopathology serving as the reference standard. Discrepancies were resolved through discussion and consensus. The quality of evidence for our pooled analyses was assessed across the patient selection, index test, reference standard, and flow and timing. Each domain was assessed for risk of bias, and the first three domains were evaluated for applicability concerns.

### 2.6. Statistical analyses

Network meta-analysis was conducted to analyze the diagnostic accuracy of 37 imaging modalities used to detect different stages of prostate cancer, as shown in Table 2, with comparison to cytological examination. For the assessment of diagnostic accuracy, pairwise analyses were conducted to estimate the odds ratio (OR) for recurrence detection and the corresponding 95% confidence interval (CI), which were calculated from sensitivity, specificity, positive predictive value, and negative predictive value in the included manuscripts.

**Table 2:** Overview of imaging modalities included in the network meta-analysis. This table provides an overview of the different imaging modalities considered in the network meta-analysis, including their abbreviations and full names.

To assist in interpreting the diagnostic performance, the Surface Under the Cumulative Ranking Curve (SUCRA) was utilized to calculate the probability of prostate cancer at various stages, which is the best diagnostic method, employing a Bayesian approach. A higher SUCRA value indicated a superior rank for the intervention. For the consistency test, node-splitting assessments were conducted to determine the association between the direct and indirect evidence. Additionally, publication bias was evaluated using funnel plots. Stata 17.0 software was utilized for data analysis in this study.

## 3. Result

### 3.1 Study selection and characteristics

The literature search yielded 24,910 references with related content. During the screening, 8,916 articles were removed due to duplication, and 1,5132 articles were excluded for irrelevance, specific screening process in Supplementary Figure 1. Of the full-text articles assessed, 739 articles were excluded based on screening criteria.

Ultimately, after screening, a total of 123 relevant articles were included. Specifically, 38 articles were included for the diagnosis of primary prostate cancer, involving 2,182 patients; 25 articles were related to lymph node metastasis, involving 1,803 patients; 41 articles were concerned with bone metastasis, involving 3,196 patients; and 37 articles were related to biochemical recurrence of prostate cancer, involving 2,190 patients.

Supplementary Figures 2 and 3 show the results regarding the risk of bias and applicability, with red dots indicating a high risk of bias for each bias criterion, yellow dots indicating an unclear risk, and green dots indicating a low risk of bias.

### 3.2 Network meta-analysis of diagnostic test accuracy for prostate cancer

#### 3.2.1 Early-phase prostate cancer

A total of 38 (23, 28–64) studies were included in this analysis. The networks of eligible comparisons are graphically represented in network plots on the diagnostic values of imaging modalities for the diagnosis of early-phase prostate cancer in Figure 1. In sensitivity comparisons, 18F-DCFBC PET/CT (Imaging14) was inferior to mpMRI (Imaging 2) (OR 0.13, 95% CI 0.02-0.75), 99mTc-PSMA SPECT/CT (Imaging 4) (OR 0.03, 95% CI 0.00-0.97), 18F-DCFPyL PET/CT (Imaging 15) (OR 0.07, 95% CI 0.01-0.61), 68Ga-P16-093 PET/CT (Imaging 29) (OR 0.01, 95% CI 0.00-0.95), and 68Ga-PSMA-11 PET/MRI (Imaging30) (OR 0.09, 95% CI 0.01-0.62). The results of the current NMA revealed that 68Ga-P16-093 PET/CT (Image29) and 68Ga-PSMA-617 PET/CT (Image33) exhibited superior diagnostic efficacy in Se, Sp, PPV and NPV, as detailed in Supplementary Figure 4.

**Figure 1:**
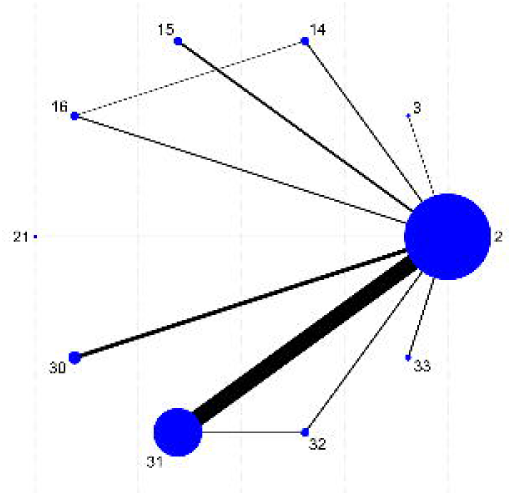
Network plots on the diagnostic values of imaging modalities for the diagnosis of early-phase prostate cancer. This figure displays the network of eligible comparisons graphically, illustrating the diagnostic values of various imaging modalities used for the detection of early-phase prostate cancer.

In specificity comparisons, 18F-DCFPyL PET/CT (Imaging 15) was inferior to 18F-DCFBC PET/CT (Imaging 14) (OR 0.07, 95% CI 0.01-0.93) and 68Ga-PSMA-617 PET/CT (Imaging 33) (OR 0.05, 95% CI 0.00-0.70), while 18F-DCFPyL PET (Imaging17) was inferior to mpMRI (Imaging 2) (OR 0.02, 95% CI 0.00-0.35), 99mTc-PSMA SPECT/CT (Imaging 4) (OR 0.02, 95% CI 0.00-0.91), 18F-DCFBC PET/CT (Imaging 14) (OR 0.00, 95% CI 0.00-0.14), 18F-DCFPyL PET/MRI (Imaging 16) (OR 0.03, 95% CI 0.00-0.46), 68Ga-PSMA-11 PET/MRI (Imaging30) (OR 0.02, 95% CI 0.00-0.36), 68Ga-PSMA-11 PET/CT (Imaging 31) (OR 0.01, 95% CI 0.00-0.25), 68Ga-PSMA-11 PET (Imaging32) (OR 0.04, 95% CI 0.00-0.67), 68Ga-PSMA-617 PET/CT (Imaging 33) (OR 0.00, 95% CI 0.00-0.09), and 68Ga-RM2 PET/CT (Imaging36) (OR 0.02, 95% CI 0.00-0.50). In positive predictive value comparisons, 18F-DCFPyL PET (Imaging 17) was inferior to mpMRI (Imaging 2) (OR 0.25, 95% CI 0.07-0.90), 68Ga-PSMA-11 PET/MRI (Imaging 30) (OR 0.19, 95% CI 0.05-0.76), 68Ga-PSMA-11 PET/CT (Imaging31) (OR 0.20, 95% CI 0.05-0.77), and 68Ga-PSMA-617 PET/CT (Imaging 33) (OR 0.06, 95% CI 0.01-0.42). 68Ga-PSMA-617 PET/CT (Imaging33) was superior to 99mTc-MDP SPECT/CT (Imaging3) (OR 68.48, 95% CI 1.35-3483.61), 18F-choline PET/CT (Imaging 11) (OR 8.70, 95% CI 1.27-59.80), 18F-DCFPyL PET/MRI (Imaging16) (OR 6.21, 95% CI 1.12-34.34), and 18F-DCFPyL PET (Imaging 17) (OR 16.60, 95% CI 2.39-115.03). There were no statistically significant differences in negative predictive value comparisons between imaging modalities and other tests, with detailed content presented in Supplementary Table 2. As such, the consistency model was applied to the current study (all p > 0.05).

In the analysis of SUCRA values for different imaging modalities, sensitivity was highest for 68Ga-P16-093 PET/CT (Imaging29) (84.1%), specificity was highest for 68Ga-PSMA-617 PET/CT (Imaging33) (88.1%), positive predictive value was highest for 68Ga-PSMA-617 PET/CT (Imaging33) (90.5%), and negative predictive value was highest for 68Ga-P16-093 PET/CT (Imaging 29) (80.4%). Detailed content is presented in Figure 2, which combines four plots.

**Figure 2:**
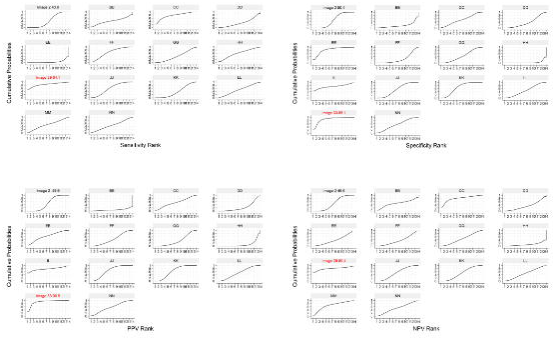
SUCRA values for different imaging modalities in early-phase prostate cancer diagnosis based on sensitivity, specificity, positive predictive value (PPV), and negative predictive value (NPV). This figure presents the Surface Under the Cumulative Ranking Curve (SUCRA) values for various imaging modalities, indicating their relative diagnostic performance in early-phase prostate cancer detection based on sensitivity, specificity, PPV, and NPV.

##### 3.2.1.1 Subgroup analysis: Peripheral invasion

In clinically significant prostate cancer, the primary outcome of nine studies was the diagnostic performance for csPCa. The results of the current NMA revealed that 18F-DCFPyL PET/CT (Image 15) and 68Ga-PSMA-11 PET/MRI (Image 30) exhibited superior diagnostic efficacy in Se, Sp, PPV and NPV, as detailed in the Supplementary Figure 5.

No significant statistical differences were observed in sensitivity comparisons among imaging modalities (P > 0.05). In specificity comparisons, 18F-DCFPyL PET (Image 17) was inferior to mpMRI (Image 2) (OR: 0.02, 95% CI: 0.00-0.22), 18F-DCFPyL PET/MRI (Image 16) (OR: 0.03, 95% CI: 0.00-0.34), 68Ga-PSMA-11 PET/MRI (Image 30) (OR: 0.01, 95% CI: 0.00-0.14), and 68Ga-PSMA-11 PET/CT (Image31) (OR: 0.02, 95% CI: 0.00-0.20). In positive predictive value (PPV), 18F-DCFPyL PET (Imaging 17) was weaker than mpMRI (Image 2) (OR: 0.24, 95% CI: 0.08-0.73), 68Ga-PSMA-11 PET/MRI (Image 30) (OR: 0.14, 95% CI: 0.03-0.71), and 68Ga-PSMA-11 PET/CT (Image31) (OR: 0.23, 95% CI: 0.07-0.71). No significant statistical differences were found in negative predictive value (NPV) among imaging modalities, as detailed in Supplementary Table 3

In the analysis of different imaging SUCRA values, 18F-DCFPyL PET/CT (Image15) had the highest sensitivity and NPV of 65.8 and 65.0, respectively, while 68Ga-PSMA-11 PET/MRI (Image 30) had the highest specificity and PPV of 85.0 and 83.7, respectively. The details are shown in Figure 3 (four graphs combined).

**Figure 3:**
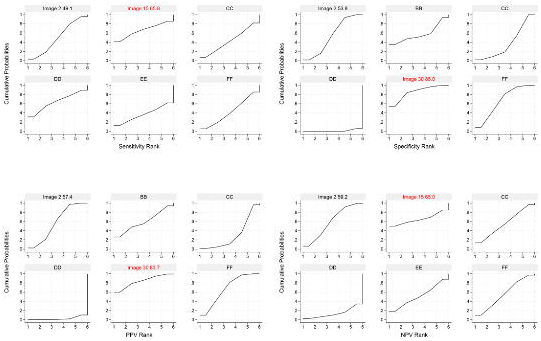
SUCRA values for different imaging modalities in clinically significant prostate cancer diagnosis based on sensitivity, specificity, positive predictive value (PPV), and negative predictive value (NPV). This figure presents the Surface Under the Cumulative Ranking Curve (SUCRA) values for various imaging modalities, indicating their relative diagnostic performance in clinically significant prostate cancer detection based on sensitivity, specificity, PPV, and NPV.

Among the studies related to extracapsular extension (ECE), a total of eight studies involved the diagnosis of ECE. The results of the current NMA revealed that mpMRI (Image 2), 18F-PSMA-1007 PET/CT (Image 21), and 68Ga-PSMA-11 PET/MRI (Image exhibited superior diagnostic efficacy in Se, Sp, PPV and NPV, as detailed in the Supplementary Figure 6. No significant statistical differences were observed in sensitivity, specificity, PPV or NPV comparisons among imaging modalities. In the SUCRA values analysis, 68Ga-PSMA-11 PET/MRI (Image 30) had the highest sensitivity and NPV of 83.0 and 78.1, respectively. 18F-PSMA-1007 PET/CT (Image 21) had the highest specificity of 78.3, while mpMRI (Image 2) had the highest PPV of 62.5. The details are shown in Figure 4 (four graphs combined).

**Figure 4:**
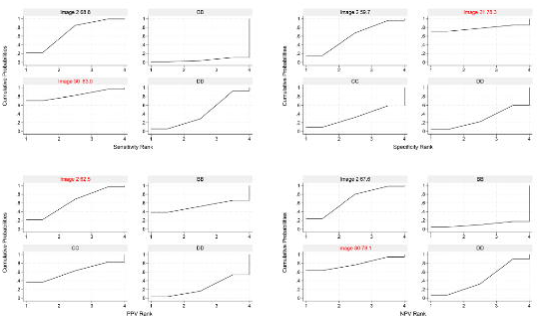
SUCRA values for different imaging modalities in extracapsular extension prostate cancer diagnosis based on sensitivity, specificity, positive predictive value (PPV), and negative predictive value (NPV). This figure presents the Surface Under the Cumulative Ranking Curve (SUCRA) values for various imaging modalities, indicating their relative diagnostic performance in extracapsular extension prostate cancer detection based on sensitivity, specificity, PPV, and NPV.

In studies related to Seminal Vesicle Invasion (SVI), the primary outcome of ten studies was the diagnosis of SVI. The results of the current NMA revealed that mpMRI (Image 2) and 18F-PSMA-1007 PET/CT (Image 21) exhibited superior diagnostic efficacy in Se, Sp, PPV and NPV, as detailed in Supplementary Figure 7.

No significant statistical differences were observed in sensitivity, specificity, PPV, or NPV comparisons among imaging modalities. In the SUCRA values analysis, 18F-PSMA-1007 PET/CT (Image 21) performed best in sensitivity, PPV, and NPV, with values of 83.3, 70.0, and 79.5, respectively. mpMRI (Image 2) performed best in specificity, with a value of 68.4. The details are shown in Figure 5 (four graphs combined).

**Figure 5:**
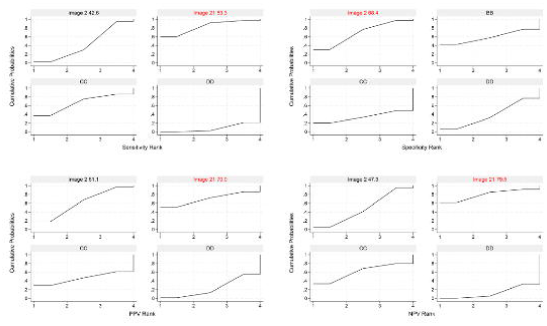
SUCRA values for different imaging modalities in seminal vesicle invasion prostate cancer diagnosis based on sensitivity, specificity, positive predictive value (PPV), and negative predictive value (NPV). This figure presents the Surface Under the Cumulative Ranking Curve (SUCRA) values for various imaging modalities, indicating their relative diagnostic performance in seminal vesicle invasion prostate cancer detection based on sensitivity, specificity, PPV, and NPV.

#### 3.2.2 lymph node metastasis

This study included 25 (28–30, 32, 46, 50, 52, 58, 59, 65–80) articles on lymph node metastasis in the initial staging of prostate cancer. The networks of eligible comparisons are graphically represented in network plots, which depict the diagnostic values of imaging examinations for the diagnosis of lymph node metastasis in prostate cancer, as shown in Figure 6. The results of the current NMA revealed that MRI (Image 1), mpMRI (Image 2), CT (Image5), 18F-DCFPyL PET/CT (Image15), 68Ga-PSMA-11 PET/MRI (Image30) and 68Ga-PSMA-11 PET/CT (Image31) showed poor diagnostic efficacy in Se, Sp, PPV and NPV, as detailed in the Supplementary Figure 8.

**Figure 6:**
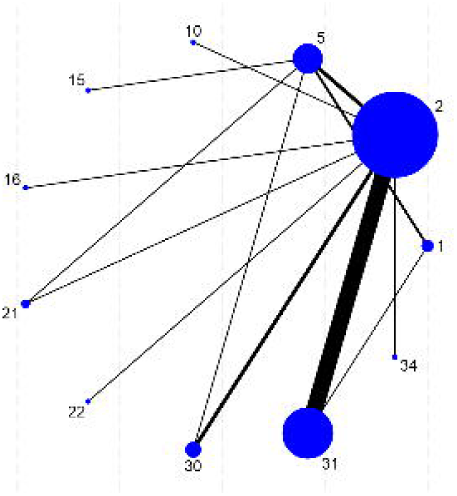
Network plots on the diagnostic values of imaging modalities for the diagnosis of lymph node metastasis prostate cancer. This figure displays the network of eligible comparisons graphically, illustrating the diagnostic values of various imaging modalities used for the detection of lymph node metastasis prostate cancer.

In the comparison of sensitivity, 68Ga-PSMA-11 PET/CT (Image 31) exhibited higher sensitivity than CT (Image 5) (OR: 4.63, 95% CI: 1.63-13.13) and mpMRI (Image 2) (OR: 3.35, 95% CI: 2.14-5.24). Additionally, 68Ga-PSMA-11 PET/CT (Image 31) demonstrated an advantage in sensitivity compared to 18F-DCFPyL PET/CT (Image 15) (OR: 4.63, 95% CI: 1.63-13.13).

However, there were no statistically significant differences in sensitivity when compared to 18F-PSMA-1007 PET/CT (Image 21), 18F-PSMA-1007 PET/MRI (Image 22), and 68Ga-PSMA-11 PET/MRI (Image 30). In the specificity analysis, 18F-DCFPyL PET/CT (Image 15) performed favorably, with higher specificity than CT (Image 5) (OR: 25.33, 95% CI: 9.00-71.25) and mpMRI (Image 2) (OR: 7.05, 95% CI: 1.86-26.63). It also showed an advantage in specificity compared to 18F-PSMA-1007 PET/CT (Image 21) (OR: 8.71, 95% CI: 1.20-63.28), 18F-PSMA-1007 PET/MRI (Image 22) (OR: 66.28, 95% CI: 2.64-1664.42), and 68Ga-PSMA-11 PET/CT (Image 31) (OR: 10.12, 95% CI: 2.6-39.18). In the comparison of positive predictive value (PPV), 18F-DCFPyL PET/CT (Image 15) outperformed 68Ga-PSMA-11 PET/CT (Image 31) (OR: 6.29, 95% CI: 1.26-31.49), CT (OR: 16.75, 95% CI: 5.33-52.69), and mpMRI (Image 2) (OR: 8.25, 95% CI: 1.71-39.93). In the comparison of negative predictive value (NPV), 68Ga-PSMA-11 PET/CT (Image 31) was stronger than CT (Image 5) (OR: 2.53, 95% CI: 1.44-4.47) and mpMRI (Image 2) (OR: 1.48, 95% CI: 1.10-1.98), with no statistically significant differences compared to other imaging methods. Detailed data are presented in Supplementary Table 4. As such, the consistency model was applied to the current study, with all p-values > 0.05.

In the analysis of SUCRA values for different imaging modalities, 68Ga-PSMA-11 PET/MRI (Image 30) had the highest sensitivity, which was 82.9. 18F-DCFPyL PET/CT (Image 15) had the highest specificity and PPV, which were 96.4 and 90.0, respectively. 18F-PSMA-1007 PET/CT (Image 21) had the highest NPV, which was 84.1. Detailed data are presented in Figure 7, which combines four plots.

**Figure 7:**
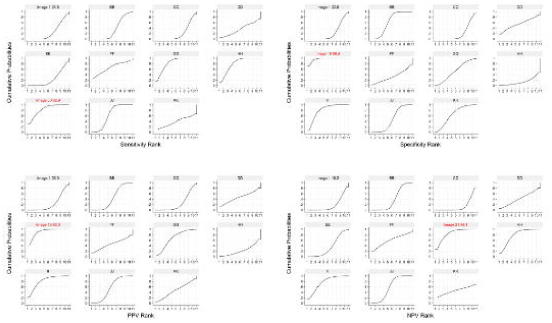
SUCRA values for different imaging modalities in lymph node metastasis prostate cancer diagnosis based on sensitivity, specificity, positive predictive value (PPV), and negative predictive value (NPV). This figure presents the Surface Under the Cumulative Ranking Curve (SUCRA) values for various imaging modalities, indicating their relative diagnostic performance in lymph node metastasis prostate cancer detection based on sensitivity, specificity, PPV, and NPV.

#### 3.2.3 bone metastasis

In the analysis of bone metastases in prostate cancer, a total of 41 (28, 73, 80–118) studies were included. The networks of eligible comparisons are graphically represented in network plots on the diagnostic values of imaging examinations for the diagnosis of bone metastases in prostate cancer in Figure 8. The results of the current NMA revealed that 99mTc-PSMA SPECT/CT (Image 4), 18F-DCFPyL PET/CT (Image 15), 18F-Fluciclovine PET/CT (Image 19), 18F-PSMA-1007 PET/CT (Image 21), 18F-PSMA-1007 PET/MRI (Image 22), 68Ga-PSMA-11 PET/MRI (Image 30) and 68Ga-PSMA-617 PET/CT (Image 33) exhibited poor diagnostic efficacy in Se, Sp, PPV and NPV, as detailed in Supplementary Figure 9.

**Figure 8:**
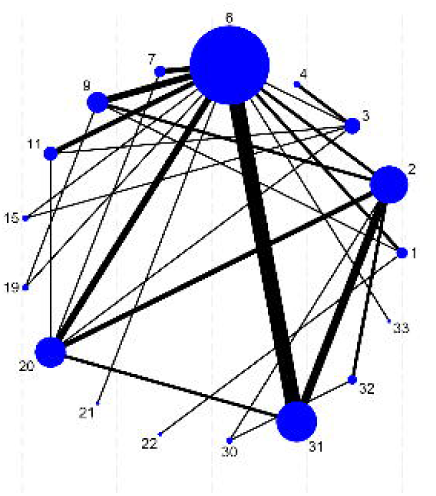
Network plots on the diagnostic values of imaging modalities for the diagnosis of bone metastasis prostate cancer. This figure displays the network of eligible comparisons graphically, illustrating the diagnostic values of various imaging modalities used for the detection of bone metastasis prostate cancer.

In the sensitivity analysis, 18F-PSMA-1007 PET/MRI (Image 22) demonstrated higher sensitivity compared to 99mTc-MDP SPECT/CT (Image 4) (OR: 58.15, 95% CI: 3.19-1058.40), BS (Image 6) (OR: 142.35, 95% CI: 11.45-1769.55), 11C-choline PET/CT (Image 9) (OR: 42.29, 95% CI: 2.95-605.66), 18F-choline PET/CT (Image 11) (OR: 39.27, 95% CI: 2.55-604.24), 18F-Fluciclovine PET/CT (Image 19) (OR: 31.61, 95% CI: 1.35-740.03), 18F-NaF PET/CT (Image 20) (OR: 18.47, 95% CI: 1.27-269.18), and 68Ga-PSMA-11 PET/MRI (Image 30) (OR: 42.67, 95% CI: 1.16-1570.95), but there were no statistically significant differences when compared to 18F-PSMA-1007 PET/CT (Image 21), 18F-DCFPyL PET/CT (Image 15), and 68Ga-PSMA-617 PET/CT (Image 33). In terms of specificity, BS (Image 6) was lower than mpMRI (Image 2) (OR: 0.31, 95% CI: 0.11-0.91), 11C-choline PET/CT (Image 9) (OR: 0.15, 95% CI: 0.04-0.50), 18F-choline PET/CT (Image 11) (OR: 0.18, 95% CI: 0.05-0.68), 18F-Fluciclovine PET/CT (Image 19) (OR: 0.10, 95% CI: 0.01-0.83), and 68Ga-PSMA-11 PET/CT (Image 31) (OR: 0.14, 95% CI: 0.06-0.31). For positive predictive value comparison, 68Ga-PSMA-11 PET/CT (Image was higher than mpMRI (Image 2) (OR: 2.69, 95% CI: 1.08-6.70), BS (Image 6) (OR: 6.84, 95% CI: 3.43-13.61), and 18F-NaF PET/CT (Image 20) (OR: 3.16, 95% CI: 1.16-8.60). In terms of negative predictive value, 18F-PSMA-1007 PET/MRI (Image 22) generally performed better and had advantages over other imaging methods, except for no statistically significant differences compared to 18F-PSMA-1007 PET/CT (Image 21), 68Ga-PSMA-11 PET/MRI (Image 30), 18F-Fluciclovine PET/CT (Image 19), 18F-DCFPyL PET/CT (Image 15), and 68Ga-PSMA-617 PET/CT (Image 33). The detailed content is shown in Supplementary Table 5. As such, the consistency model was applied to the current study (all p > 0.05).

In the SUCRA ranking plot, 18F-PSMA-1007 PET/MRI (Image 22) showed the highest sensitivity and negative predictive value of 97.4% and 95.9%, respectively. For specificity and positive predictive value, 18F-Fluciclovine PET/CT (Image 19) had the highest SUCRA values of 74.2% and 76.3%, respectively. The detailed content is shown in Figure 9 (four plots together).

**Figure 9:**
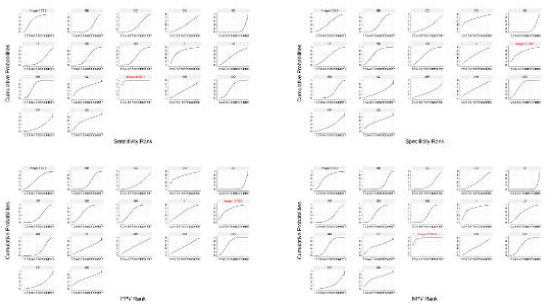
SUCRA values for different imaging modalities in bone metastasis prostate cancer diagnosis based on sensitivity, specificity, positive predictive value (PPV), and negative predictive value (NPV). This figure presents the Surface Under the Cumulative Ranking Curve (SUCRA) values for various imaging modalities, indicating their relative diagnostic performance in bone metastasis prostate cancer detection based on sensitivity, specificity, PPV, and NPV.

##### 3.2.3.1 Subgroup Analysis Newly Diagnosed and Biochemically Recurrent Prostate Cancer

The studies included in the bone metastasis analysis were categorized into two groups based on disease stage: newly diagnosed and biochemically recurrent prostate cancer. The remaining studies involved a mixed analysis of biochemical recurrence and newly diagnosed and were not included in the subgroup analysis. Additionally, due to the unknown initial treatment types and androgen deprivation therapy (ADT) proportions in the studies involving biochemical recurrence patients, relevant subgroup analysis could not be conducted.

In newly diagnosed prostate cancer bone metastases, a total of 17 studies were included. The results of the current NMA revealed that mpMRI (Image 2), 18F-DCFPyL PET/CT (Image 15), 18F-PSMA-1007 PET/MRI (Image 22), 68Ga-PSMA-11 PET/MRI (Image 30), 68Ga-PSMA-11 PET/CT (Image 31), 68Ga-PSMA-11 PET (Image 32) and 68Ga-PSMA-617 PET/CT (Image 33) exhibited poor diagnostic efficacy in Se, Sp, PPV and NPV, as detailed in Supplementary Figure 10. For sensitivity comparisons, the sensitivity of BS (Image 6) was lower than that of 18F-choline PET/CT (Image 11) (OR: 0.20, 95% CI: 0.06-0.68), 18F-DCFPyL PET/CT (Image 15) (OR: 0.06, 95% CI: 0.01-0.72), 18F-NaF PET/CT (Image 20) (OR: 0.13, 95% CI: 0.04-0.42), 18F-PSMA-1007 PET/MRI (Image 22) (OR: 0.03, 95% CI: 0.00-0.61), and 68Ga-PSMA-11 PET/CT (Image 31) (OR: 0.11, 95% CI: 0.03-0.47). However, there was no statistical difference compared to mpMRI (Image 2) (OR: 0.09, 95% CI: 0.01-1.16) and 99mTc-MDP SPECT/CT (Image 3) (OR: 0.33, 95% CI: 0.06-1.90). In terms of specificity, BS (Image 6) specificity was lower than that of mpMRI (Image 2) (OR: 0.13, 95% CI: 0.02-0.89), 99mTc-MDP SPECT (Image 7) (OR: 0.08, 95% CI: 0.01-0.53), and 68Ga-PSMA-11 PET/CT (Image 31) (OR: 0.14, 95% CI: 0.04-0.45). Similar to specificity, BS (Image 6) was lower than mpMRI (Image 2), 99mTc-MDP SPECT (Image 7), and 68Ga-PSMA-11 PET/CT (Image 31) in positive predictive value. When comparing negative predictive values, BS (Image 6) was lower than other imaging methods except for 68Ga-PSMA-617 PET/CT (Image 33), mpMRI (Image 2), and 99mTc-MDP SPECT/CT (Image 3). Specific details are shown in Supplementary Table 6. As such, the consistency model was applied to the current study (all p > 0.05).

In the analysis of SUCRA values for different imaging techniques, 68Ga-PSMA-11 PET (Image 32) showed the best performance in sensitivity and negative predictive value, with values of 86.6% and 85.9%, respectively. 99mTc-MDP SPECT (Image 3) had the highest specificity of 82.3%. mpMRI (Image 2) showed the highest positive predictive value of 82.1%. Specific details are shown in Figure 10.

**Figure 10:**
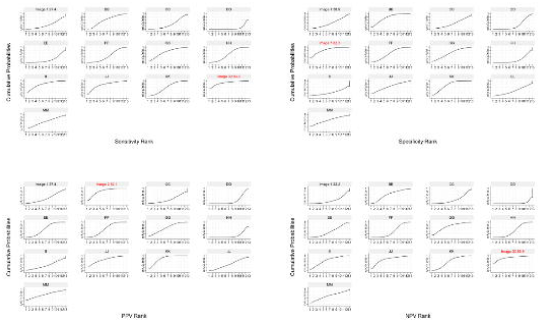
SUCRA values of different imaging modalities based on sensitivity, specificity, positive predictive value (PPV) and negative predictive value (NPV) in the diagnosis of bone metastases in newly diagnosed prostate cancer. This figure presents the Surface Under the Cumulative Ranking Curve (SUCRA) values for various imaging modalities, indicating their relative diagnostic performance in the detection of bone metastases in newly diagnosed prostate cancer based on sensitivity, specificity, PPV, and NPV.

In biochemically recurrent prostate cancer bone metastases, a total of 17 studies were included. The results of the current NMA revealed that mpMRI (Image 2), BS (Image 6), 11C-choline PET/CT (Image 9) and 68Ga-PSMA-11 PET/CT (Image 31) exhibited poor diagnostic efficacy in Se, Sp, PPV and NPV, as detailed in the Supplementary Figure 11. No statistical difference was found in sensitivity and negative predictive value comparisons between imaging examinations. In terms of specificity, BS (Image 6) was lower than mpMRI (Image 2) (OR: 0.20, 95% CI: 0.06-0.70), 11C-choline PET/CT (Image 9) (OR: 0.10, 95% CI: 0.03-0.31), and 68Ga-PSMA-11 PET/CT (Image 31) (OR: 0.16, 95% CI: 0.06-0.44). In positive predictive value comparisons, BS (Image 6) was lower than 11C-choline PET/CT (Image 9) (OR: 0.13, 95% CI: 0.04-0.41) and 68Ga-PSMA-11 PET/CT (Image 31) (OR: 0.17, 95% CI: 0.06-0.48), with no significant statistical difference compared to other imaging methods. Specific details are shown in Supplementary Table 7. As such, the consistency model was applied to the current study (all p > 0.05).

In the SUCRA ranking chart, 18F-NaF PET/CT (Image 20) showed the highest sensitivity of 79.5%. In terms of specificity, 68Ga-PSMA-11 PET (Image 32) had the highest SUCRA value of 83.8%. For positive predictive value and negative predictive value, 18F-Fluciclovine PET/CT (Image 19) and MRI (Image 1) performed best, with values of 78.6% and 85.1%, respectively. Specific details are shown in Figure 11.

**Figure 11:**
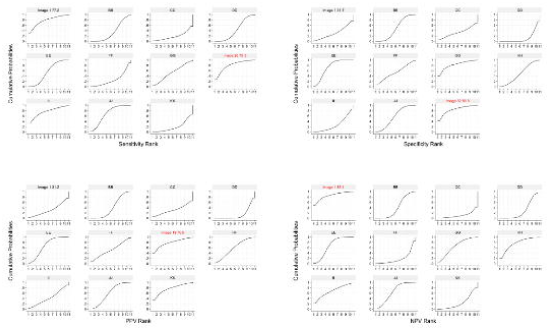
SUCRA values of different imaging modalities based on sensitivity, specificity, positive predictive value (PPV) and negative predictive value (NPV) in the diagnosis of biochemically recurrent prostate cancer bone metastases. This figure presents the Surface Under the Cumulative Ranking Curve (SUCRA) values for various imaging modalities, indicating their relative diagnostic performance in the detection of biochemically recurrent prostate cancer bone metastases based on sensitivity, specificity, PPV, and NPV.

#### 3.2.4 biochemical recurrence

In the investigation of biochemical recurrence, a total of 37 (28, 102, 108–110, 112, 119–149) studies were included. The networks of eligible comparisons are graphically represented in network plots, showcasing the diagnostic values of imaging examinations for the diagnosis of biochemical recurrence of prostate cancer in Figure 12. Notably, 68Ga-NeoB PET/MRI (Image 28) and 68Ga-PSMA-R2 PET/MRI (Image 35) did not form network relationships with other diagnostic methods and were therefore excluded from subsequent analyses. The results of the current NMA revealed that mpMRI (Image 2), BS (Image 6), 11C-choline PET/CT (Image 9) and 68Ga-PSMA-11 PET/CT (Image 31) were poor in terms of lesion detection rates, as detailed in the Supplementary Table 8. Ranked by SUCRA values, 18F-PSMA-1007 PET/CT (Image 21) exhibited the best performance in detecting biochemical recurrence, with a rate of 98.1. The specific details are illustrated in Figure 13.

**Figure 12:**
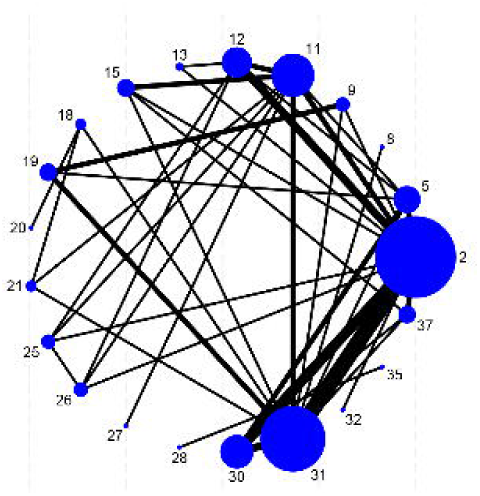
Network plots on the diagnostic values of imaging modalities for the diagnosis of biochemical recurrence cancer. This figure displays the network of eligible comparisons graphically, illustrating the diagnostic values of various imaging modalities used for the detection of biochemical recurrence of prostate cancer.

**Figure 13:**
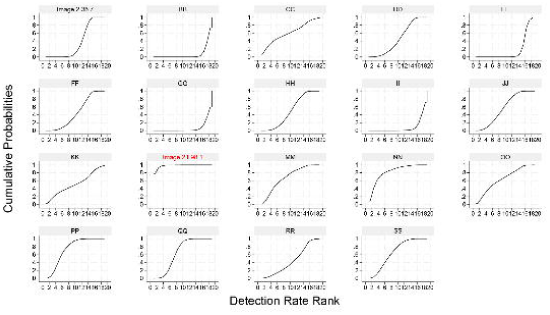
SUCRA values for different imaging modalities in biochemical recurrence of prostate cancer diagnosis based on detection rate (DR). This figure presents the Surface Under the Cumulative Ranking Curve (SUCRA) values for various imaging modalities, indicating their relative diagnostic performance in biochemical recurrence of prostate cancer detection based on detection rate (DR).

##### 3.2.4.1 Subgroup Analysis: Radioactive Markers

Furthermore, this study conducted an additional analysis and comparison of the detection rates of biochemical recurrence in prostate cancer, classified according to radioactive tracers. Notably, two radioactive tracers, 68Ga-PSMA-R2 and 68Ga-NeoB, did not form network relationships with other tracers and were therefore excluded from subsequent analyses. The estimated scanning rate ratios for pairwise comparisons among the radioactive markers in the NMA are presented in Supplementary Table 9. Ranked by SUCRA values, 18F-PSMA-1007 exhibited the best performance in detecting biochemical recurrence, with a rate of 98.8. The detection rates of 64Cu-PSMA-617, 68Ga-PSMA-11, and 18F-DCFPyL were comparable, with rates of 74.9, 70.0, and 66.3, respectively. The specific details are illustrated in Figure 14.

**Figure 14:**
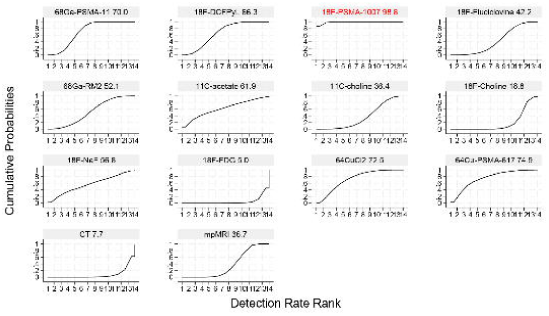
SUCRA values for different radiotracers in the diagnosis of biochemical recurrence of prostate cancer based on detection rate (DR). This figure presents the Surface Under the Cumulative Ranking Curve (SUCRA) values for various radiotracers, indicating their relative diagnostic performance in biochemical recurrence of prostate cancer detection based on detection rate (DR).

### 3.3 Bias analysis: Small sample effect estimation

Funnel plots were drawn for the total effective outcome indicator to test for publication bias. The results showed that all studies were generally symmetrically distributed around the X = 0 vertical line, and most studies fell inside the funnel, whereas some fell at the bottom, suggesting a possible small sample effect (Figure 15).

**Figure 15:**
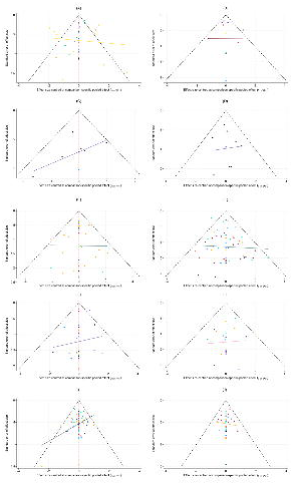
Funnel plots of various imaging modalities for different stages of prostate cancer. (A) Funnel plot of early prostate cancer; (B) Funnel plot of clinically significant prostate cancer; (C) Funnel plot of extracapsular extension; (D) Funnel plot of seminal vesicle invasion; (E) Funnel plot of lymph node metastasis; (F) Funnel plot of bone metastasis; (G) Funnel plot of bone metastases in newly diagnosed prostate cancer; (H) Funnel plot of bone metastases in biochemically recurrent prostate cancer; (I) Funnel plot of biochemical recurrence of prostate cancer; (J) Funnel plot of biochemically recurrent prostate cancer with different radiotracers.

## 4. Discussion

In this systematic review and network meta-analysis, we comprehensively evaluated the diagnostic efficacy of various imaging modalities for prostate cancer at different stages, including early-phase, lymph node metastasis, bone metastasis, and biochemical recurrence. Our findings highlight the strengths and limitations of each imaging technique in detecting and staging prostate cancer.

The 5-year survival rate of prostate cancer is highly correlated with tumor staging, and improving the accuracy of early diagnosis can significantly enhance patient survival rates (150). In the early diagnosis of prostate cancer, mpMRI is recommended before biopsy, and patients with PI-RADS ≤ 2 may be exempted from prostate cancer biopsy (151). Prostate-specific membrane antigen (PSMA) is a type 2 transmembrane glycoprotein that provides necessary metabolic substrates for cancer cell proliferation and invasion by mediating folate hydrolysis (152). In high-grade metastatic prostate cancer, particularly in aggressive, androgen-deprived, metastatic, and hormone-refractory PCa, PSMA expression is significantly increased and is currently the main target for detecting minimal prostate cancer lesions (153, 154). PSMA mediates folate hydrolysis, providing essential metabolic substrates for cancer cell proliferation and invasion.

PET/CT tracers primarily encompass three functionalities: detecting cell division activity (such as radiolabeled choline, acetate, fluorodeoxyglucose, amino acids), targeting cancer-specific membrane proteins or receptors (like prostate-specific membrane antigen, gastrin-releasing peptide receptors), and compounds specifically binding to bone metastases (e.g., radiolabeled sodium fluoride) (155). Due to PSMA’s involvement in the PI3K/AKT growth pathway related to prostate cancer metastasis (156), PSMA-PET can better reflect the overall tumor burden in the body (157).

The PSMA/PET evaluation system integrates PSMA expression V (based on background contrast) and PSMA expression Q (using SUV values), where significantly higher PSMA expression than the liver is considered a typical pathological feature of prostate cancer (158). This system demonstrates higher SUVmax values in more aggressive tumors, further confirming its diagnostic efficacy (159). Multi-Parametric MRI (mpMRI) combines anatomical imaging (such as T2-weighted MRI, which differentiates anatomical structures based on water molecule relaxation time differences and possesses excellent spatial resolution) with functional imaging (including Diffusion-Weighted Imaging (DWI) to assess water molecule diffusion in tissue structures, where tumor regions appear as high signals; and DCE-MRI to evaluate angiogenesis and perfusion characteristics through contrast agent injection, providing information on tumor blood supply) (160). Studies have shown that the combined use of mpMRI and PSMA PET/CT significantly improves the negative predictive value and sensitivity of diagnosis (31), which may be related to the targeted biopsy capabilities of PSMA PET/CT for patients with metal implants. In the imaging diagnosis of prostate cancer, MRI-TRUS fusion imaging has become a clinically preferred option (7).

Establishing a high-precision diagnostic method for early-stage prostate cancer is crucial for reducing unnecessary biopsies and the associated risks of overtreatment and invasive injuries to patients. Current research indicates that Digital Rectal Examination (DRE) performs poorly in early screening for prostate cancer, potentially increasing the risk of overtreatment and unnecessary physical harm (11, 161). Particularly in middle-aged men, the diagnostic efficacy of DRE is inferior to PSA screening. Therefore, screening and diagnostic strategies for prostate cancer should comprehensively consider patients’ expected lifespan and health status. According to the EAU-EANM-ESTRO-ESUR-ISUP-SIOG guidelines, local treatment is recommended for patients with an expected lifespan exceeding ten years, while those with a shorter lifespan (e.g., <10 to 15 years) may be more suitable for active surveillance or androgen deprivation therapy (162).

The accuracy of mpMRI (multiparametric magnetic resonance imaging) in early diagnosis of prostate cancer is highly dependent on the professional expertise of imaging specialists. However, with the deep integration of computer science and medical imaging technology, particularly the application of advanced computer-aided diagnosis (CAD) techniques such as Convolutional Neural Networks (CNNs), the precision of mpMRI in the early detection of prostate cancer has been significantly enhanced (163). This advancement provides strong technical support for the early and precise diagnosis of prostate cancer.

In advanced cases, bone metastasis is one of the leading causes of death (164). Planar bone scintigraphy (BS) using Technetium-99m (99mTc) diphosphonates is currently a primary imaging modality for assessing high-risk prostate cancer, characterized by low cost, high sensitivity, and low specificity (165). Bone activation induced by hormonal therapy can result in heterogeneous bone uptake, masking bone metastasis(166). False positives mainly occur in noncancerous bone conditions of the spine and ribs (91).

The axial skeleton is the primary site of skeletal-related events (SREs) in prostate cancer (167). In the diagnosis of advanced disease, PSMA PET/CT demonstrates higher accuracy, lower radiation exposure, and fewer equivocal diagnostic lesions compared to traditional imaging modalities such as CT and bone scans (168), as well as higher interobserver agreement (169). In the selection of molecular hybrid imaging ligands, 18F-PSMA-1007 is primarily eliminated via the hepatobiliary route, while 68Ga-PSMA-11 is excreted through both the hepatobiliary and urinary systems; thus 18F-PSMA-1007 exhibits better diagnostic performance (170), especially in lesions around the bladder (171), unspecific bone uptake (UBU) (172), and soft tissue surrounding the lesion (90). Delayed imaging can be used to increase diagnostic accuracy in patients with high bladder urine activity (173).

In the assessment of single bone metastasis lesions, 18F-NaF PET/CT demonstrates superiority over 68Ga-PSMA PET/CT (174). This is attributed to its longer half-life (175) and higher interobserver agreement (176). Compared to PET/CT, PET/MRI exhibits greater sensitivity in the early diagnosis of bone metastases, albeit with a higher economic burden (177). Muehlematter et al. found that PSMA-PET/MRI is more sensitive than mpMRI in detecting extracapsular extension and seminal vesicle invasion in prostate cancer (45). PSMA PET also reveals the expression of PSMA in tumor-related metastatic lesions, providing a basis for potential PSMA radioligand therapy (178). Studies have also shown that 99mTc-PSMA SPECT/CT outperforms 99mTc (Methylene Diphosphonate)-MDP SPECT/CT in diagnosing bone metastases in patients with small lesions or low PSA levels. Due to differences in PSA diagnostic thresholds (99mTc-PSMA SPECT/CT: 2.635 ng/mL; 99mTc-MDP SPECT/CT: 15.275 ng/mL) and late imaging, 99mTc-PSMA SPECT/CT remains the preferred choice for patients with low PSA levels (115, 179). In assessing metastatic prostate cancer (mPCa), PSMA PET/CT defines significant increases in PSMA uptake or a >30% increase in tumor PET volume as criteria for disease progression, effectively curbing overtreatment of non-clearly progressive prostate cancer (180).

After primary treatment, biochemical recurrence (BCR) of prostate Cancer may occur, which can be classified as a negative or fossa-confined disease, lymph nodal, or distant metastatic disease. The three-year recurrence-free rate gradually decreases among these patients (181). Accurate tumor localization is crucial for guiding subsequent salvage therapies (175). Approximately 40% of patients may experience biochemical recurrence within five years after definitive treatment (182). Among patients with biochemical recurrence, 24%-34% will develop overt metastatic disease within 15 years after surgery (183). BCR is defined as a rise in PSA levels to ≥0.2 ng/ml in patients treated with radical prostatectomy or an increase in PSA to ≥2 ng/ml above the nadir in the case of primary radiotherapy, in accordance with the Phoenix criteria(184). In some cases of recurrent prostate cancer within the prostatic fossa, diagnostic accuracy can be improved by the administration of furosemide (185).

When detecting lymph node metastasis in radical prostatectomy, PSMA PET/CT exhibits high sensitivity and specificity for lymph nodes larger than 5mm (186). The primary manifestation of prostate cancer recurrence after radical prostatectomy is the presence of early enhancement foci confined to the prostatic fossa (fossa-confined disease). Post-treatment, the Prostate Imaging Reporting and Data System (PI-RADS) is no longer applicable, regardless of whether the prostate cancer was treated surgically or with primary therapy (187). Diffusion-weighted imaging-Magnetic Resonance Imaging (DWI-MRI) is the most accurate sequence for detecting prostate cancer recurrence (188). When diagnosing BCR, PSMA exhibits higher sensitivity and specificity compared to 18F-choline and 18F-fluciclovine, which rely heavily on PSA levels (123, 189).

The primary site of prostate cancer recurrence is at the bladder-ureter anastomosis, followed by the anterior or posterior bladder neck. In the diagnosis of local recurrence, PET-MRI demonstrates better recognition than PET-CT (190). 18F-PSMA-1007 PET/CT is highly specific for diagnosing lymph node recurrence after radical prostatectomy (RP), with a specificity of up to 99% (176, 191). Radiocomposites, such as 18F-NOTA-GRPR-PSMA, may represent a future direction for enhancing diagnostic efficacy (191). PSMA-PET exhibits high sensitivity for patients with high PSA levels, high Gleason scores, and rapid PSA doubling time (PSA-DT) (192). As PSMA PET/CT can be influenced by the androgen signaling pathway, it is typically recommended to wait at least three months after initiating ADT or second-generation anti-androgen therapy before performing a PSMA PET/CT scan (193).

This network meta-analysis offers a comprehensive evaluation of the diagnostic efficacy of diverse imaging modalities in the detection and staging of prostate cancer across various stages, including early-phase disease, lymph node metastasis, bone metastasis, and biochemical recurrence. Our findings emphatically highlight the strengths and limitations of each imaging technique in the context of prostate cancer detection and staging. However, due to limitations in medical and economic resources, a subset of patients remains unable to access more precise imaging modalities. As a result, high-precision examinations are often reserved for patients with advanced or overtly progressing diseases.

This study is also subject to several limitations. Firstly, the utilization of varying reference standards for different aspects of the study, along with the acquisition of some data through follow-up, may potentially impact the research findings. Secondly, the existence of potential publication bias, whereby some negative results may remain unpublished due to their non-significance, could also influence the research outcomes. Furthermore, the fact that some studies did not directly report raw data, with some data instead derived through calculations, may introduce discrepancies compared to the original data. Additionally, factors such as varying ethnic groups, different clinical scenarios, and the absence of long-term follow-up for emerging technologies can also exert an impact on the results. Finally, this study analyzed the efficacy of specific imaging methods for the diagnosis of prostate cancer. In the included literature, certain imaging methods were less well documented or involved a smaller total number of patients, and there may have been some bias when they were compared with each other. Consequently, further exploration of this issue necessitates multi-center, large-sample studies to gain a deeper understanding.

## 5. Conclusion

Our study contributes to the guidance of imaging modalities for prostate cancer across various stages. By utilizing appropriate imaging techniques, we have mitigated the potential harm caused by invasive procedures to patients. Additionally, we have provided cost-effective imaging methods for managing potential multi-stage prostate cancer conditions, thereby reducing the financial burden on patients during their medical journey. With the continuous development of ligands, precise diagnosis for an even wider range of prostate cancer stages, at a more granular level, will become a reality in the future.

## Abbreviations

ADT: Androgen Deprivation Therapy
BS: Bone Scintigraphy
BCR: Biochemical Recurrence
CAD: Computer-Aided Diagnosis
CI: Confidence Interval
CNN: Convolutional Neural Network
csPCa: Clinically Significant Prostate Cancer
CT: Computed Tomography
DCE-MRI: Dynamic Contrast-Enhanced MRI
DCFPyL: Piflufolastat
DRE: Digital Rectal Examination
DR: Detection Rate
DTA: Diagnostic Test Accuracy
DWI: Diffusion-Weighted Imaging
EAU: European Association of Urology
EANM: European Association of Nuclear Medicine
ESTRO: European Society for Radiotherapy & Oncology
ESUR: European Society of Urogenital Radiology
ECE: Extracapsular Extension
FDG: Fluoro-2-deoxy-D-glucose
FN: False Negative
FP: False Positive
ISUP: International Society of Urological Pathology
miTNM: Molecular Imaging Standardized Evaluation TNM Classification
mpMRI: Multiparametric Magnetic Resonance Imaging
MRSI: Magnetic Resonance Spectroscopy Imaging
MRE: Magnetic Resonance Elastography
NMA: Network Meta-analysis
NPV: Negative Predictive Value
OR: Odds Ratio
PCa: Prostate Cancer
PET: Positron Emission Tomography
PET/CT: Positron Emission Tomography/Computed Tomography
PET/MRI: Positron Emission Tomography/Magnetic Resonance Imaging
PI-RADS: Prostate Imaging Reporting and Data System
PPV: positive predictive value
PSA: Prostate-Specific Antigen
PSMA: Prostate-Specific Membrane Antigen
QUADAS-2: Quality Assessment of Diagnostic Accuracy Studies
RP: Radical Prostatectomy
Se: Sensitivity
Sp: Specificity
SPECT: Single Photon Emission Computed Tomography
SREs: Skeletal-Related Events
SUCRA: Surface Under Cumulative Ranking Curve
SUV: Standardized Uptake Value
SUVmax: Maximum Standardized Uptake Value
SVI: Seminal Vesicle Invasion
TP: True Positive
TN: True Negative
UBU: Unspecific Bone Uptake

**Appendix**

## Data availability statement

The original contributions presented in the study are included in the article/Supplementary Material. Further inquiries can be directed to the corresponding author.

## Acknowledgements

Thanks for all the patients in this research, and thanks for all the scholars in this article. Thanks for all the teammates for supporting this research. This work was financially supported by Research project of the Science and Technology Development Project of Jilin Province, China (20200201315JC). We are also particularly grateful to our colleagues in The First Affiliated Hospital of Jilin University for their contributions.

## Author contributions

CD S and KY: Study design, literature search and manuscript writing. YH and YT W: Study selection and data analysis. CD S and KY: Data collection. WG W and JL: Article Guidance. All authors revised the manuscript, approved the final manuscript as submitted, and agreed to take responsibility for all aspects of the work.

## Publisher’s note

All claims expressed in this article are solely those of the authors and do not necessarily represent those of their affiliated organizations or those of the publisher, the editors and the reviewers. Any product that may be evaluated in this article or claim that may be made by its manufacturer is not guaranteed or endorsed by the publisher.

