## Supplementary figures and images for "Diagnostic Efficacy of Various Imaging Modalities Across Different Stages of Prostate Cancer: A Network Meta-Analysis of Diagnostic Studies"

### Supplementary Figure 1

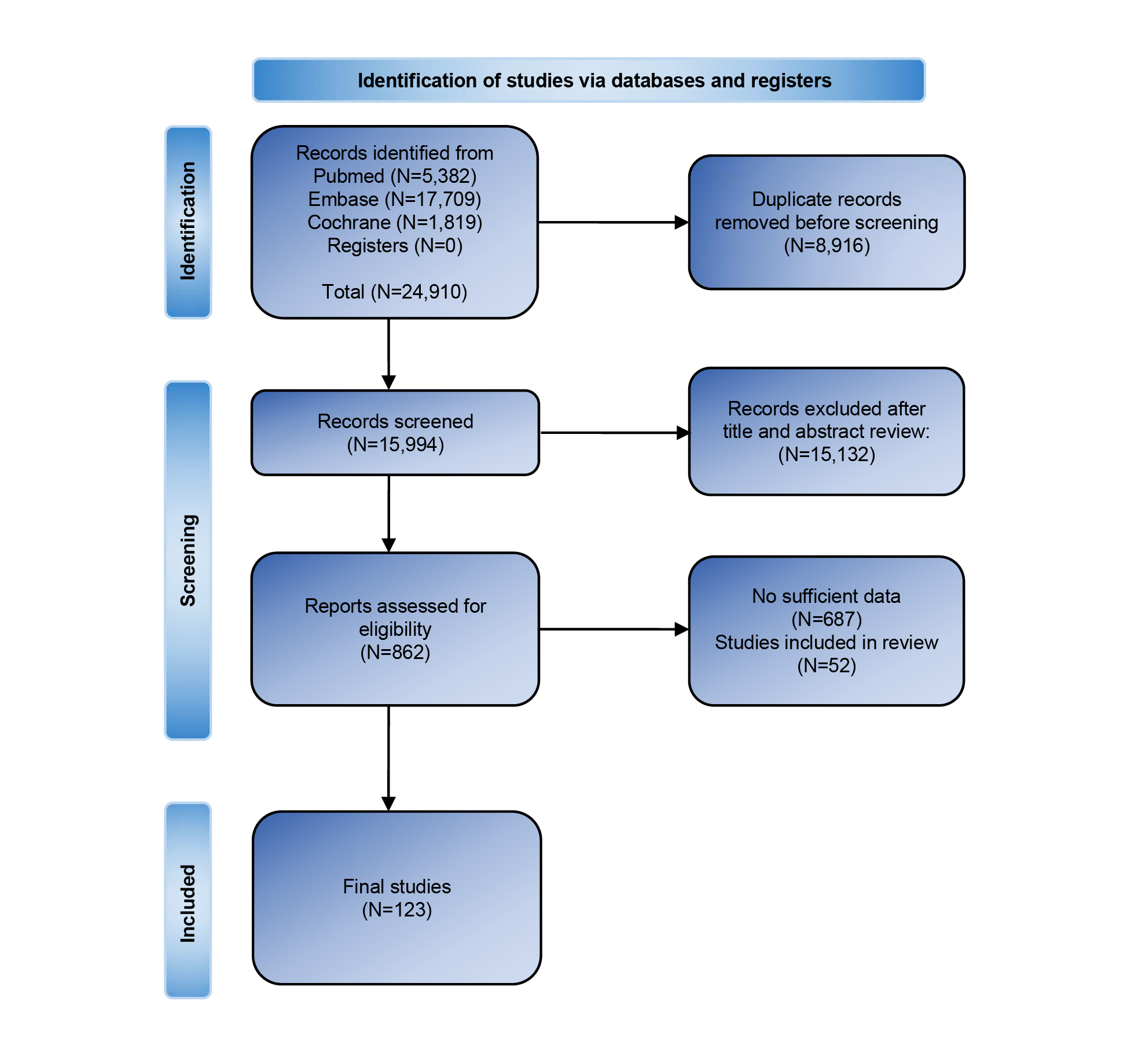

### Supplementary Figure 2

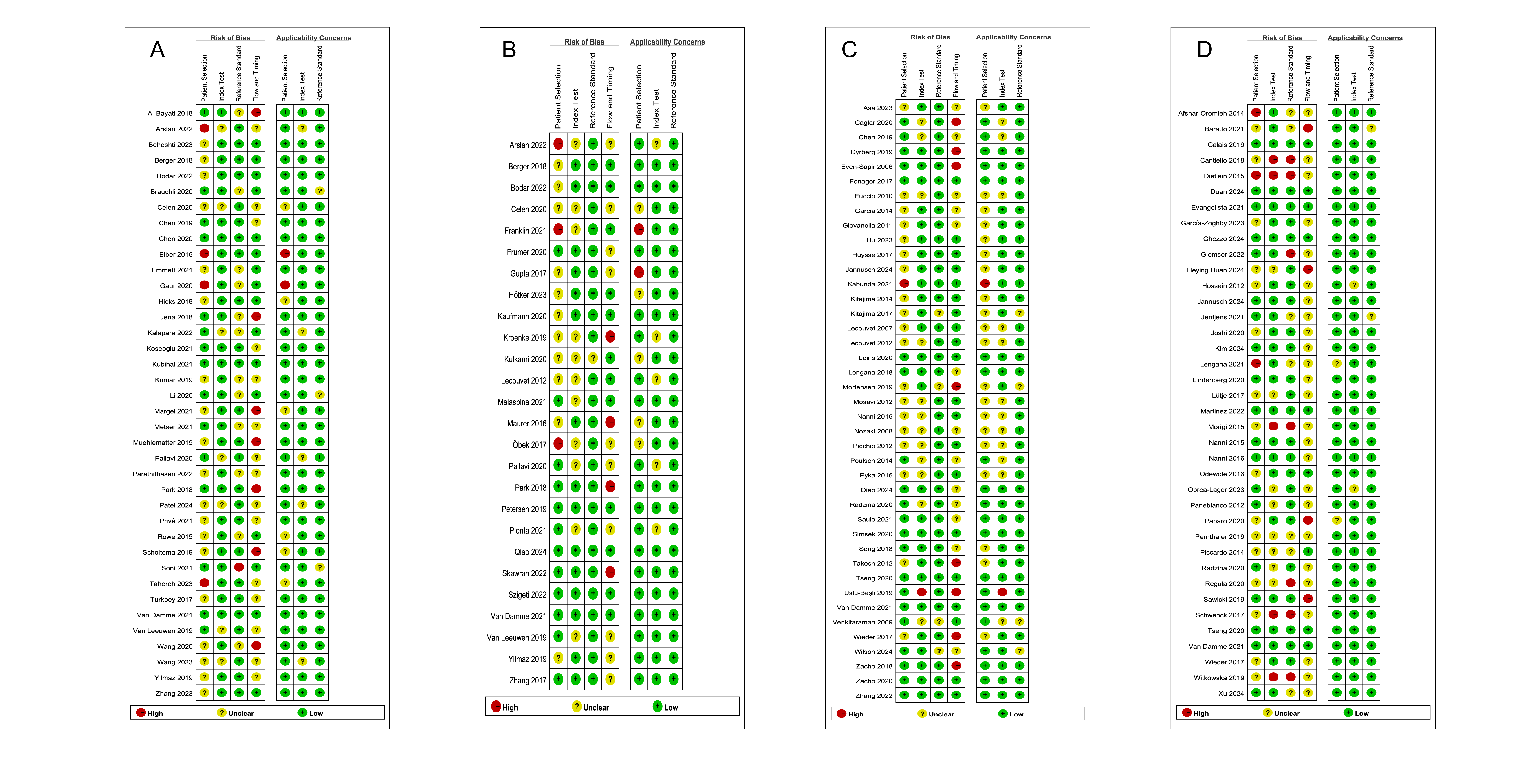

### Supplementary Figure 3

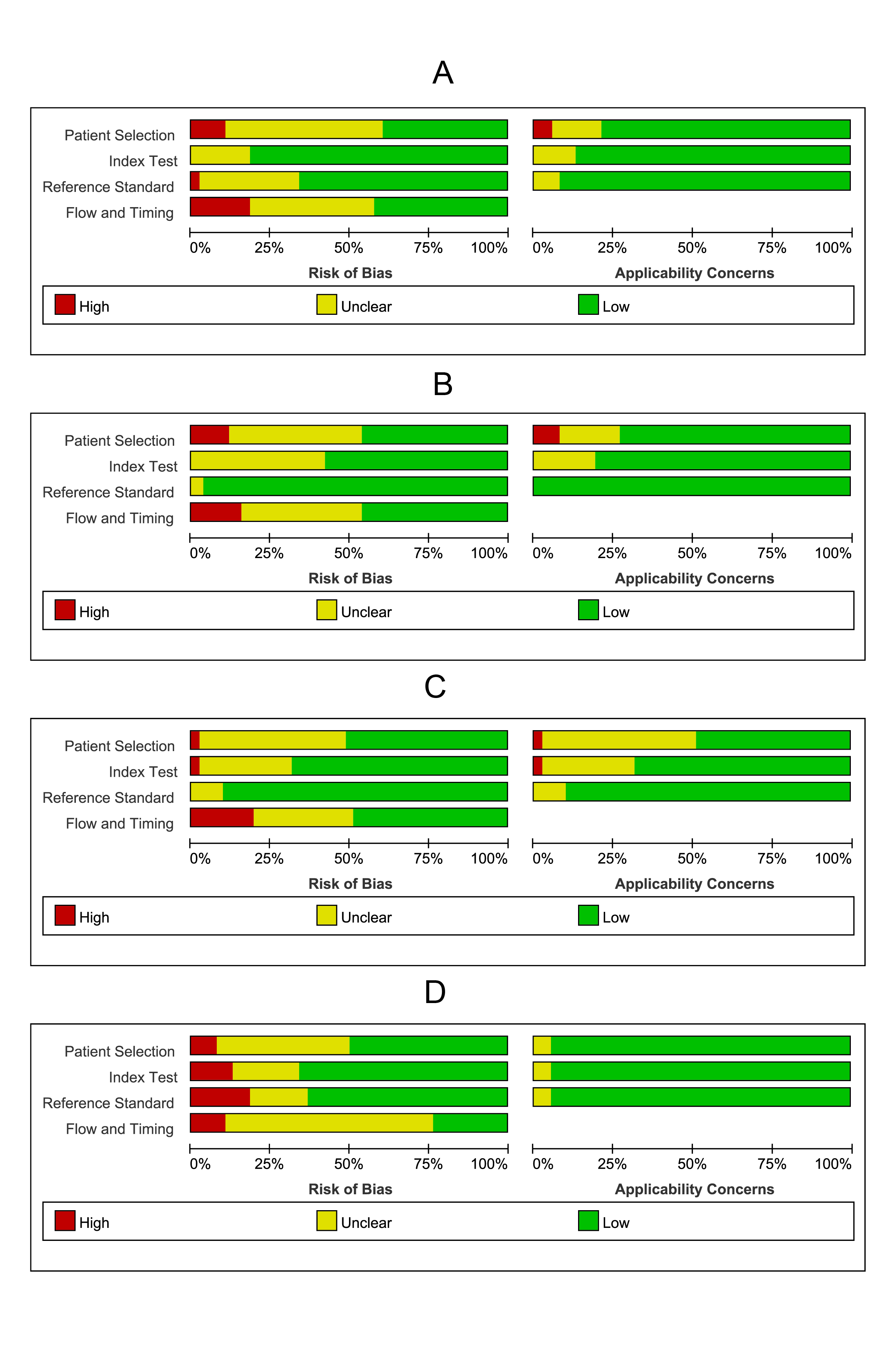

### Supplementary Figure 4

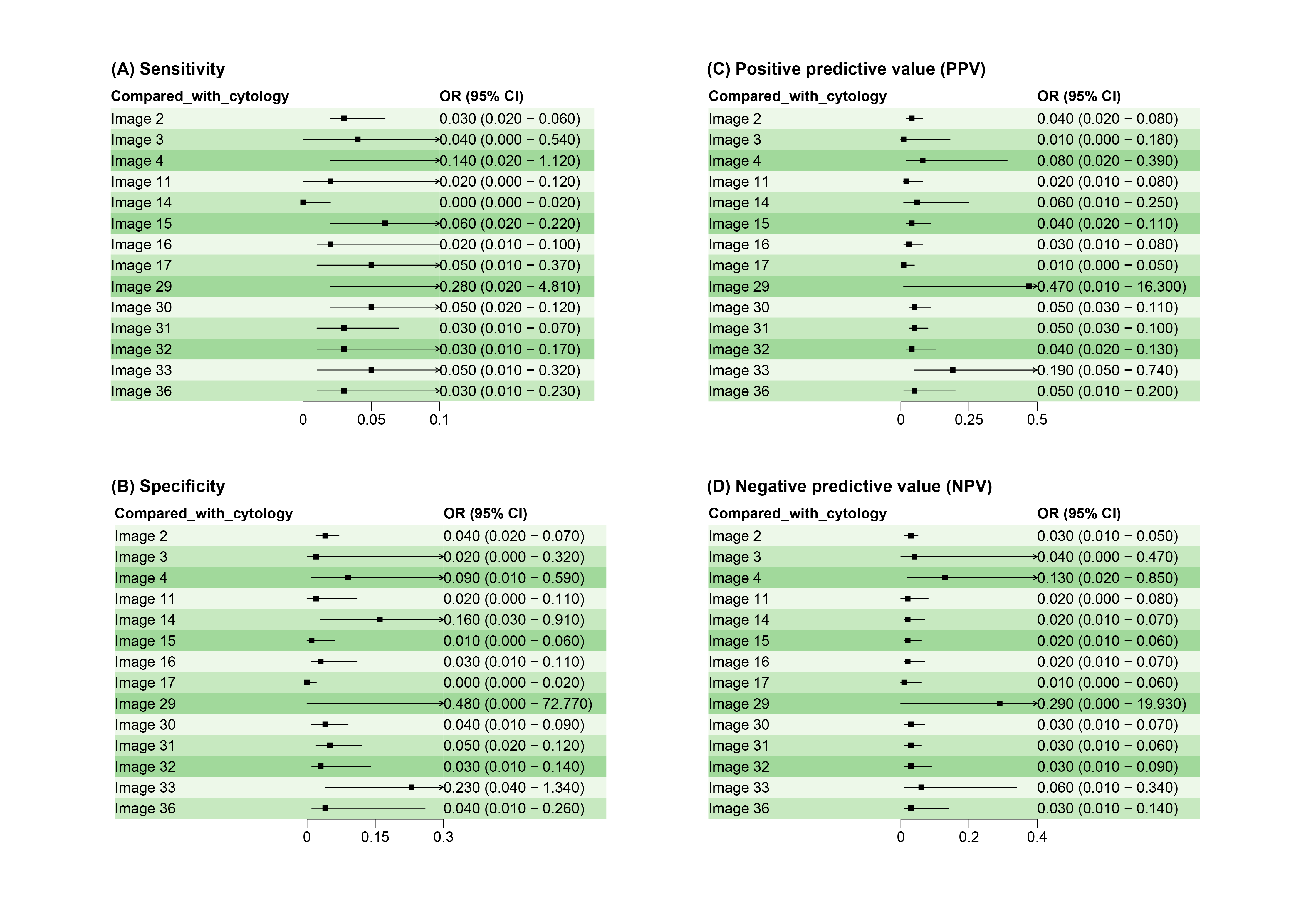

### Supplementary Figure 5

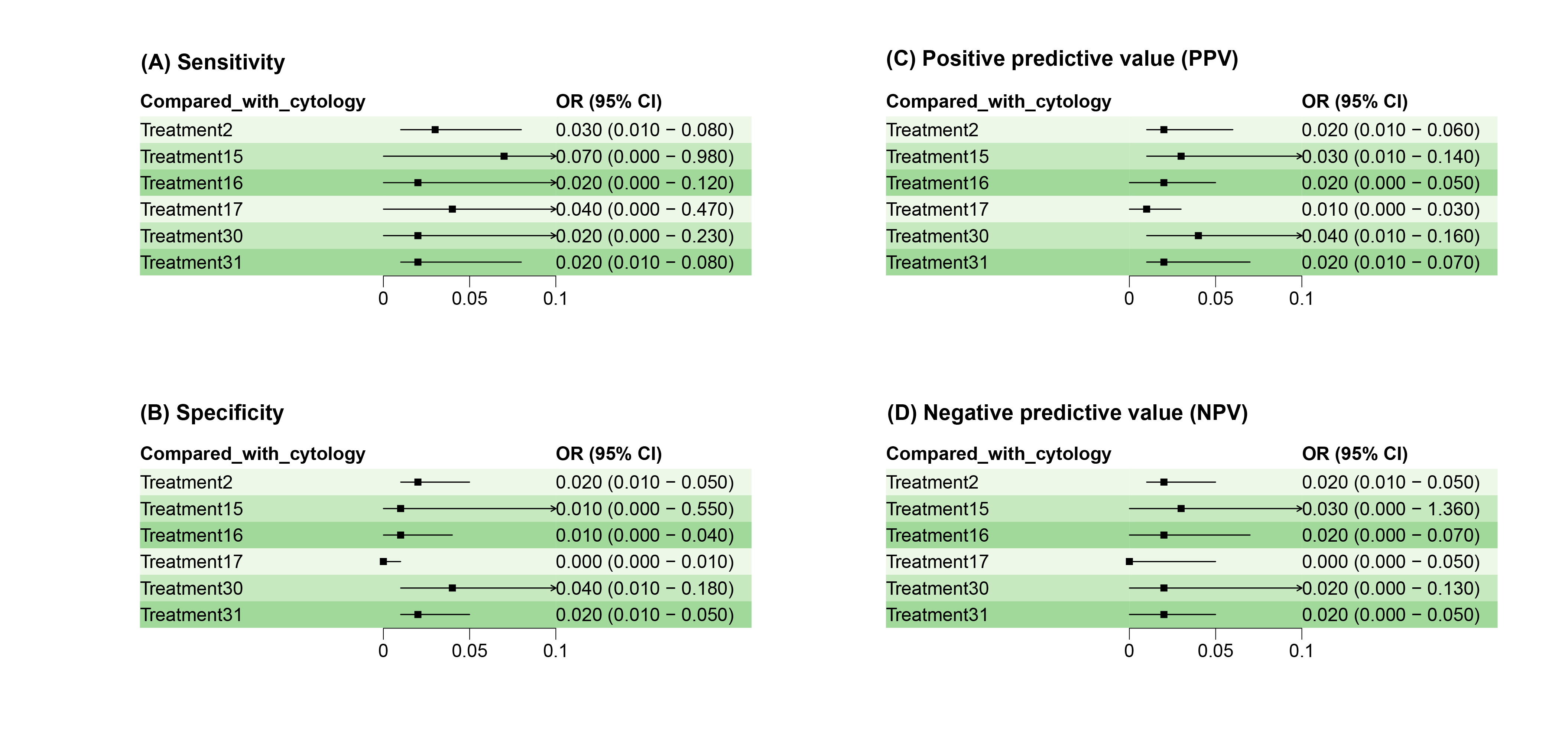

### Supplementary Figure 6

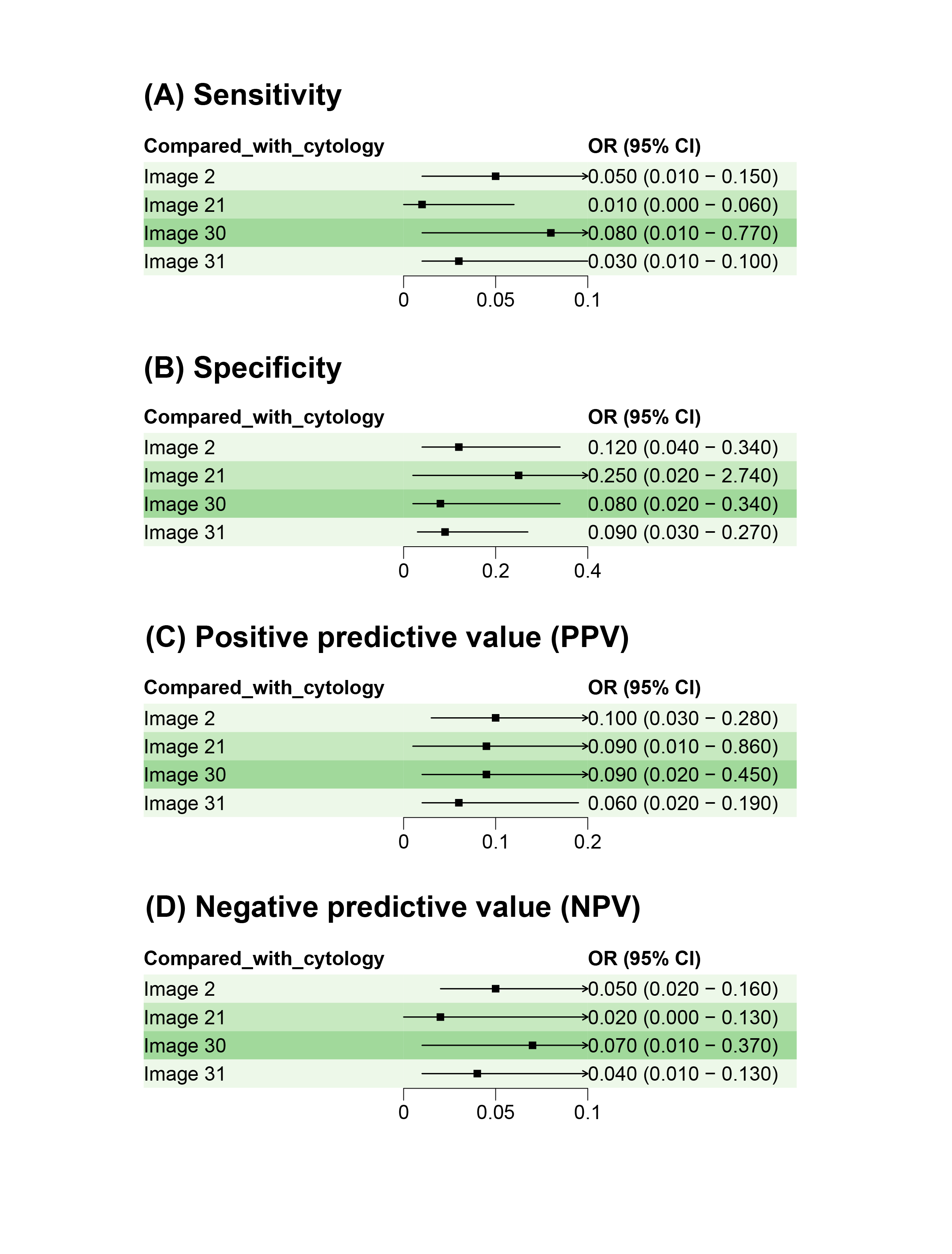

### Supplementary Figure 7

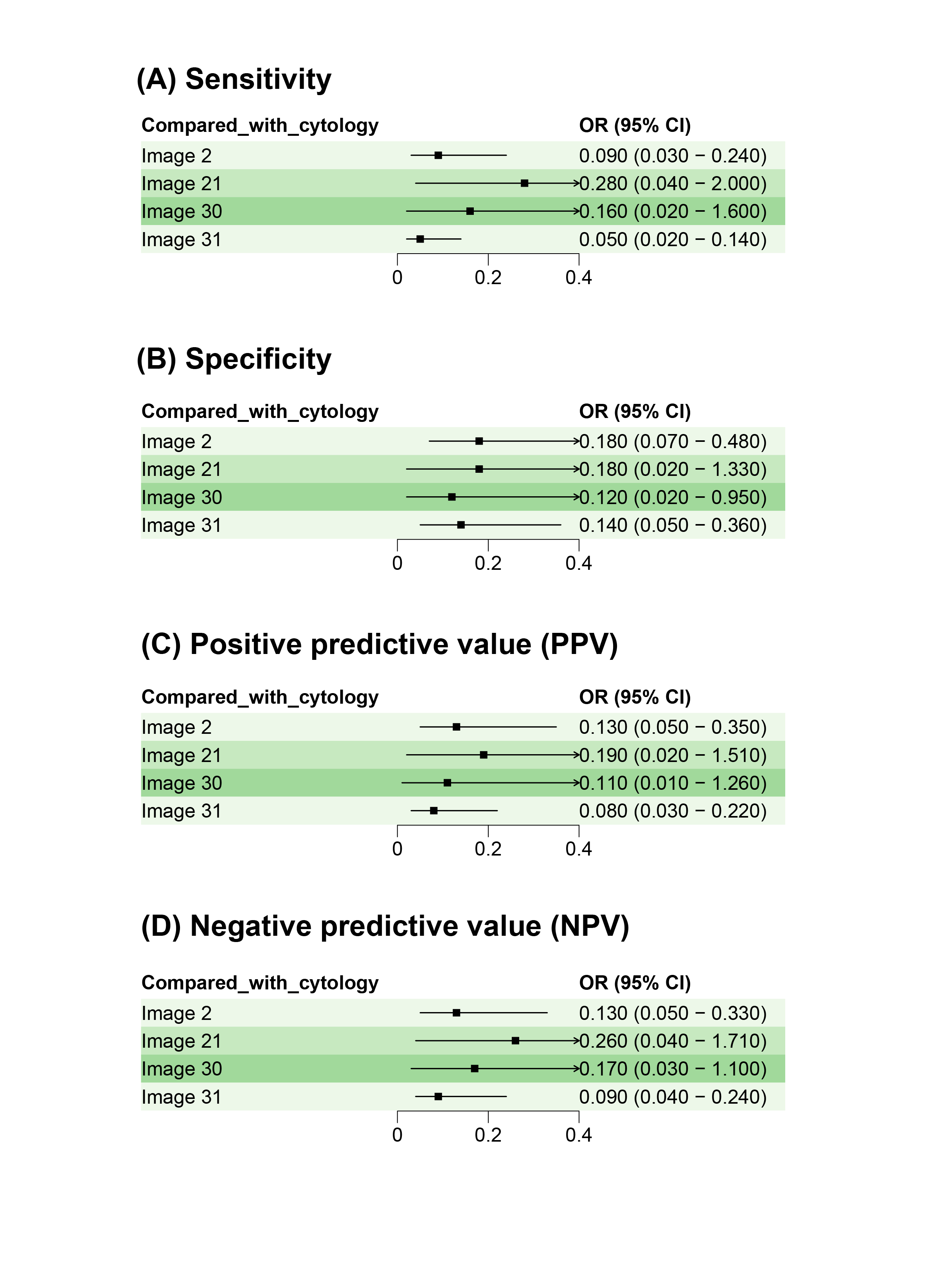

### Supplementary Figure 8

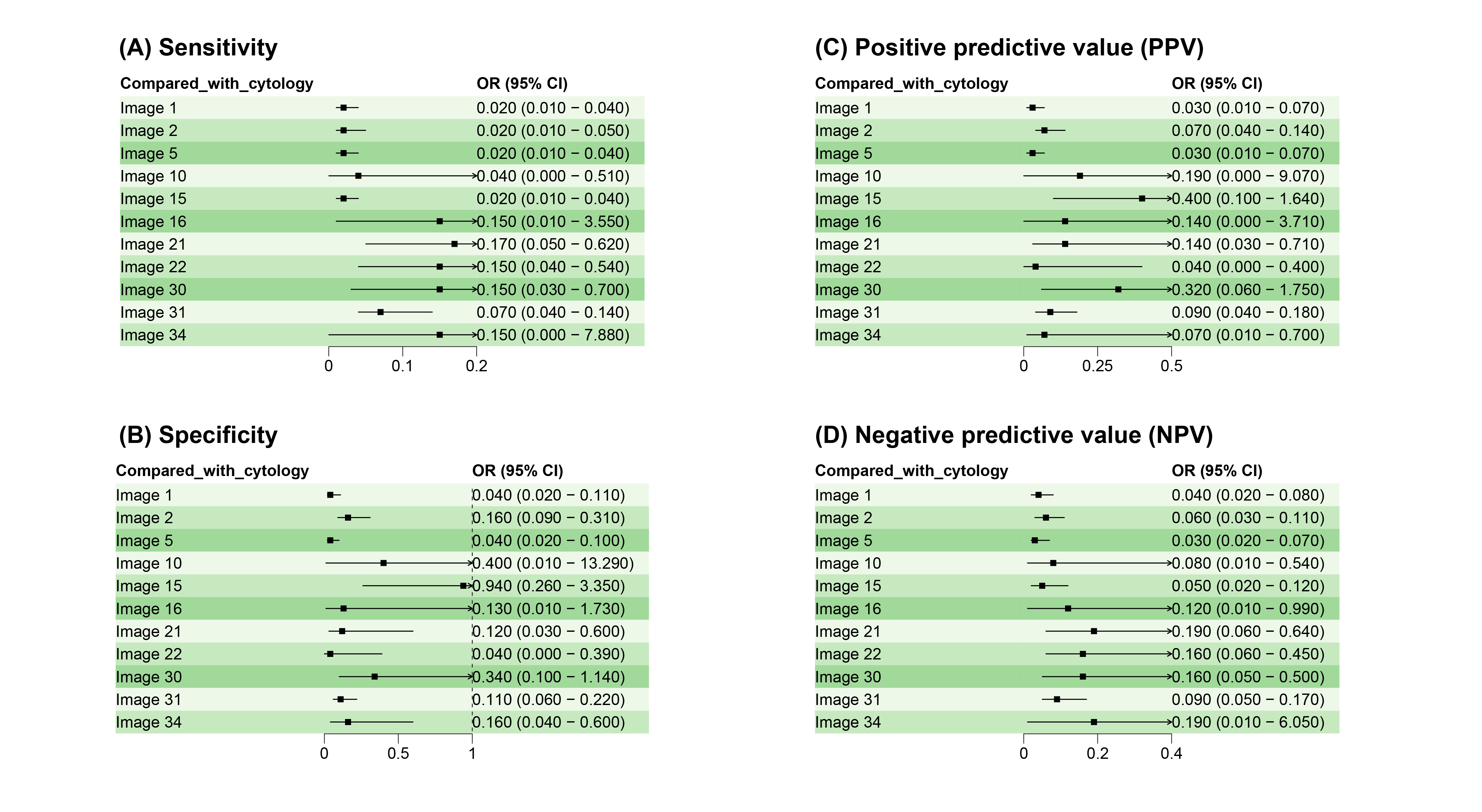

### Supplementary Figure 9

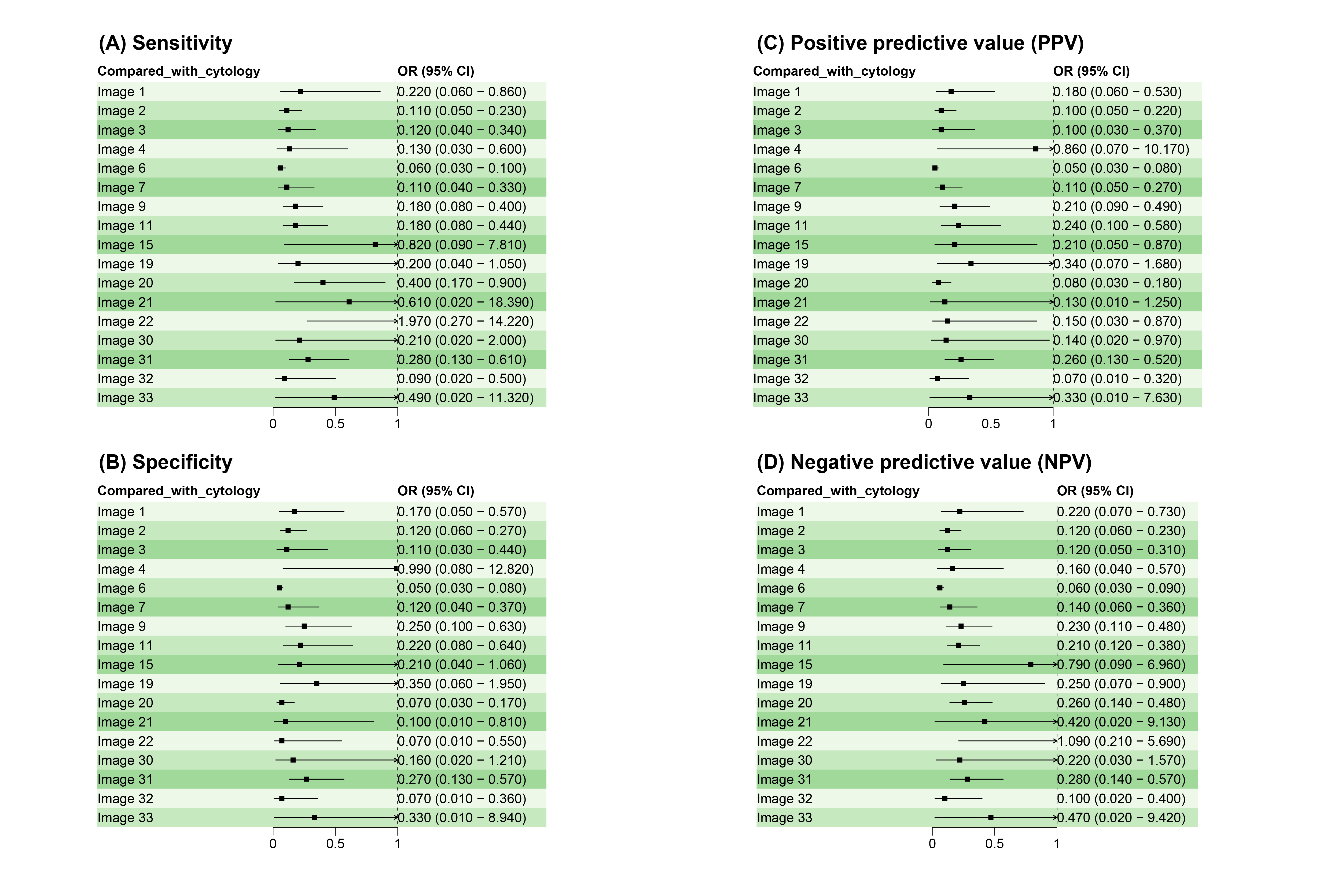

### Supplementary Figure 10

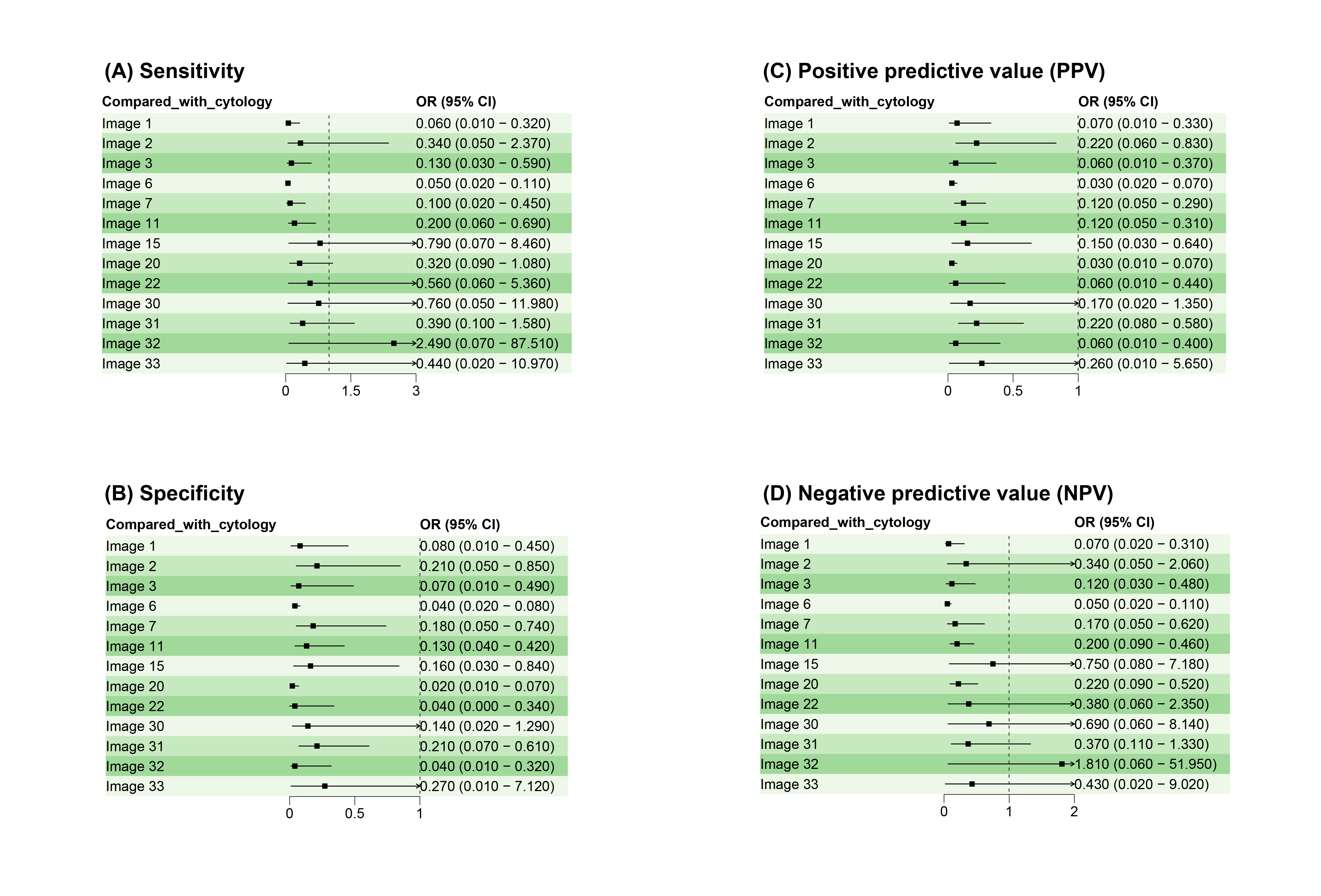

### Supplementary Figure 11

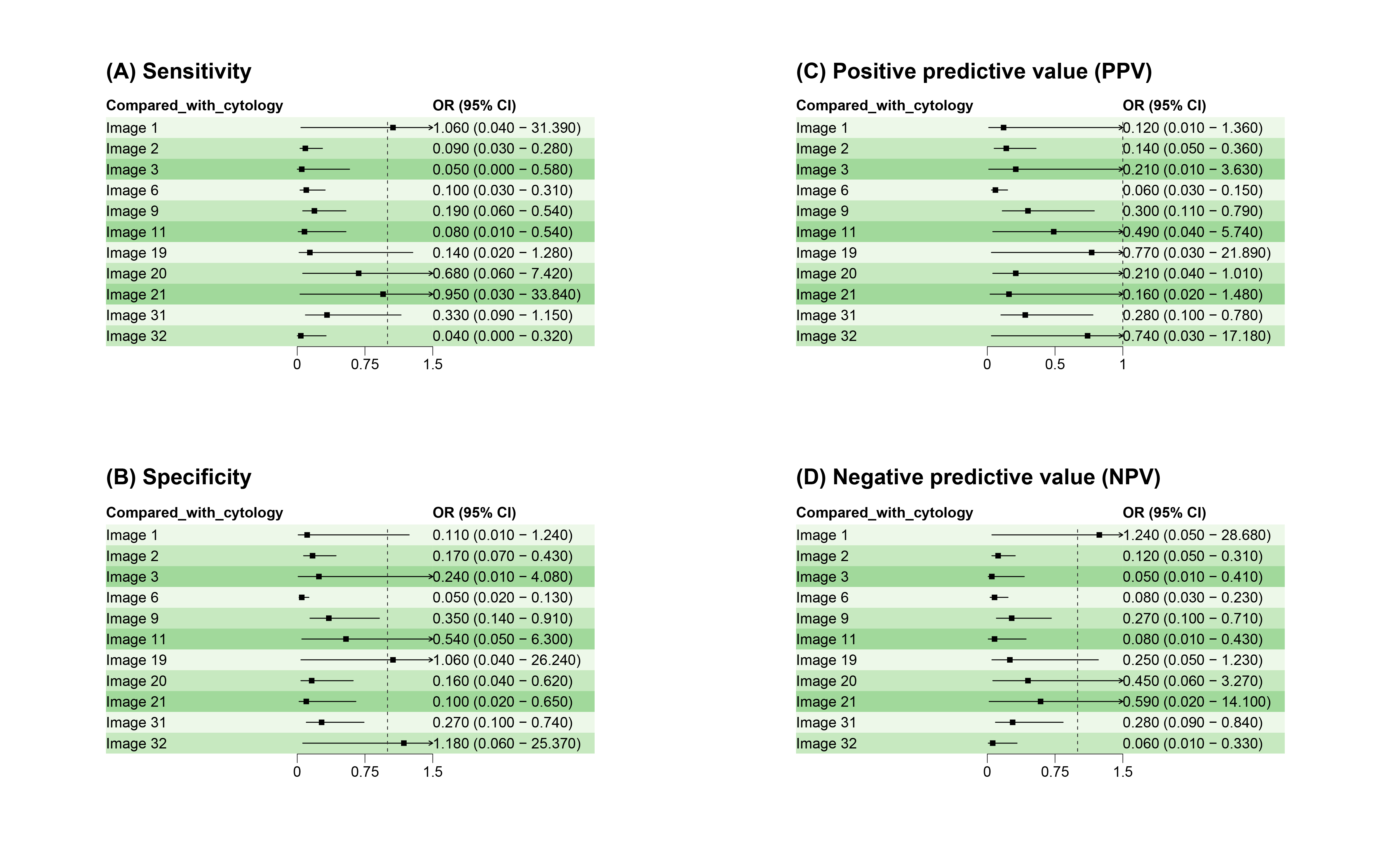
